# Communication without dialogue? Framing patient–professional interaction in UK institutional discourse on pregnancy loss

**DOI:** 10.64898/2026.11.05.26367267

**Authors:** Louise Nuttall, Beth Malory

**Affiliations:** Department of English Language and Literature, University College London, Foster Court, Gower Street, London, WC1E 6BT, UK

**Keywords:** Health communication, Patient-professional interaction, maternity care, pregnancy loss, medical organisations, charities, communicative agency

## Abstract

Communication failures are repeatedly implicated in harmful experiences and outcomes in pregnancy care, especially in contexts of loss, yet little is known about how communication itself is represented in the institutional texts that shape expectations of care. This study examines the extent to which communication is discussed, and how patient-healthcare professional interaction is framed, in publicly accessible UK discourse about pregnancy loss. Two corpora comprising texts from seven medical organisations (120,383 words) and eight charities (224,481 words) were divided into healthcare professional-facing and patient-facing sub-corpora. Frequency, semantic keyness, keyword and concordance analyses were conducted using Wmatrix7.

In medical discourse, language and communicative interaction were underrepresented, while speech acts associated with unidirectional professional activities such as advising, recommending and referring were overrepresented. Compared with charity texts, medical outputs foregrounded clinical evidence, policy, risk and professional knowledge. Characteristic forms including “confirmed”, “advised”, “recommended” and “offered” frequently occurred in passive or agentless constructions that positioned patients principally as recipients of information; patient agency was most visible in “reporting” symptoms or concerns. Charity discourse more often foregrounded talking, listening, feelings, preferences, wording, lived experience, and patient knowledge; representing communication as active and bidirectional.

We argue that UK medical discourse constructs communication primarily as the transmission of authorised knowledge rather than as relational clinical practice, reinforcing a paternalistic distribution of communicative agency. Medical institutions should embed listening, reciprocal dialogue, sensitivity to language preferences, and checks on understanding as core standards of safe pregnancy care.

## 1. Introduction

Recent decades have seen increasing acknowledgment of the impact of language and communication on patient experience across healthcare contexts (Arora, 2003; Slade et al 2008; Street et al., 2009). In pregnancy care, and especially contexts involving the loss of a baby during pregnancy, efficient and sensitive communication has been repeatedly highlighted as contributing to the experiences of individuals and families, as well as their longer-term recovery and mental health (Lacci-Reilly et al., 2023). Despite recommendations for communication in this context (e.g. Johnson et al., 2020; RCOG, 2022), detrimental impacts of poor communication around pregnancy loss continue to be highlighted internationally, both by clinicians (e.g. Kraus, 2022; Vimalesvaren et al., 2021) and members of the public (e.g. Peanut, 2023). In a recent empirical study, qualitative data from people with lived experience of pregnancy loss showed that poor communication contributes to a lack of perceived clarity, compassion, and transparency in the care received, and can exacerbate psychological trauma (Malory, 2025; Malory & Parr, 2025).

That communication is a significant factor in determining patient *experience* in this context has been recognised by the outputs of the UK’s National Maternity and Neonatal Services Investigation (MNSI), led by Baroness Valerie Amos, which concluded in 2026. The MNSI’s Interim Report (Feb 2026) highlighted communication failures including lack of sensitivity and discriminatory language, and emphasised the importance of compassionate care, which can “make the difference between leaving hospital traumatised or reassured and supported” (1). In addition to determining patient experience, MSNI outputs add to a growing body of evidence from previous UK maternity services reviews (see Malory et al., forthcoming for a summary), which indicate that communication failures in maternity care contribute to adverse pregnancy outcomes. The MNSI’s Final Report and Recommendations (June 2026) lists four core ‘concerns’ with maternity care in England; the first of these is “women and birthing people not being listened to, heard or believed” (9). Communication failures are thus foregrounded as having “serious consequences for the safety and quality of the care [women and birthing people] receive, resulting in avoidable harm, trauma and loss of confidence” (9). As such, a core recommendation of the MNSI is to “[t]reat listening to, hearing and acting on the voices of women and birthing people as a *critical safety issue*” (16; emphasis added).

Reviews therefore confirm repeatedly that the quality and consistency of communication determines both *experiences* and *outcomes* of patients across the maternity pathway, including: the extent to which families feel listened to and believed, their involvement in decision making, the way in which their concerns are responded to, and the preservation of dignity for them and their baby in contexts of loss. These issues around communication may stem from a wider lack of resources, and a specific lack of communication training for healthcare professionals involved in maternity care.

The Ockenden Report on Nottingham University Hospitals Trust’s maternity services, published in 2026, controversially indicated that a ‘toxic culture’ can underpin dismissive practices around how patients are informed, consulted and listened to (Ockenden, 2026). Framed in this way, concerns about communication which focus on individual interpersonal failures may risk alienating healthcare professionals, and undermining workforce morale. It may therefore be more helpful to consider communicative practices which seem to enact ‘medical misogyny’ as manifestations of what Malory (2027) calls “the paternalistic tendency for health *services* to undermine the testimonies of women and birthing people” (emphasis added). That dismissal of women and birthing people’s voices and perspectives and a ‘failure to listen’ recur consistently in the maternity reviews published in recent decades prompts the question: how is patient-healthcare professional communication understood and perceived in perinatal care contexts?

One way to begin to answer this question is by looking at how communication between patients and professionals is currently represented in contemporary texts about pregnancy loss produced by organisations such as the NHS, and other influential public health institutions in the UK. Employing purpose-built datasets, or *corpora*, comprising large numbers of publicly accessible texts on this subject, we use computer software to identify recurring, or statistically salient, language choices – an approach known as corpus linguistics (McEnery and Hardie, 2012). We combine this with qualitative analysis drawing on concepts from critical discourse analysis (CDA) and communication studies, such as *framing* (Entman, 1993), in order to identify the understandings and perceptions that these linguistic patterns could reflect and reinforce in this context. This combination of corpus linguistic and CDA approaches has previously been shown to offer a ‘useful methodological synergy’ through which to explore specific discourses, including in health communication contexts (Baker et al. 2008; see also Baker et al. 2020; Partington et al. 2013).

This article therefore adopts a corpus-informed, critical linguistic approach to investigating how communication between healthcare professionals and patients is framed in institutional discourses relating to pregnancy loss in the UK. Two corpora were built which aimed to be representative of the publicly accessible institutional language of a) UK medical organisations; and b) UK-based charities, specific to the topic of loss during pregnancy. These corpora were then analysed in terms of the following research questions:

1. To what extent is health communication (including language) discussed within these institutional discourses?
2. How is communication between patients and healthcare professionals framed?

## 2. Background

This research is situated in the context of the growing body of work on health communication in applied linguistics and related fields. As defined by Harvey and Adolphs (2011: 470), health communication covers ‘a huge and diverse range of communicative activities touching on health and healthcare, ranging from personal accounts of health and illness and encounters with medical professionals through to health policy documentation and side effects information presented on drug packaging’. This article examines written language used by prominent UK medical institutions and charities on their websites as examples of health discourse. We examine these discourses from a critical standpoint, drawing on the CDA understanding of ‘discourse’ as a form of social practice which not only reflects underlying attitudes, or ideologies, but also serves to construct and reinforce them (Brookes, 2026: 323-4; Fairclough, 2003; Jones, 2013). In this view, discourse ‘constitutes a central activity to the practices of institutions related to health’ (Brookes and Hunt 2021: 4).

As will be described in more detail in the next section, the discourses considered for the purpose of the present research are made up of written texts produced by UK organisations which a) share a common focus on pregnancy loss, and which b) are easily accessible to the public on their websites. They can thus be compared with other forms of public health communication such as care guidelines (Pattison, 2006), charity reports (Abnett et al., 2025) and public health campaigns (Brookes et al., 2016). Such texts are predicted to have implications for their target audiences in terms of the perceptions they construct or reinforce. Public-facing health information texts may influence patients’ expectations of, and engagement with, healthcare through their representation of health conditions and patients themselves (Brookes et al., 2016; Stanley et al., 2025). At the same time, public health communication texts also represent healthcare practitioners and the institutions in which they work. This representation can influence professional identities and self-concepts of healthcare practitioners, including understandings of how they should communicate with each other and with patients (Pattison, 2006; Sun and Chalupnik, 2022). This article explores how both patients and practitioners are represented in the institutional discourses in question, specifically in terms of the roles they take in communication, and considers the implications of these representations for different target audiences.

Before outlining the data in the following section, we first introduce a useful theoretical concept underpinning this research. The notion of ‘framing’ has been used differently in various fields including sociology, semantics, artificial intelligence and cognitive linguistics, but might broadly be defined as an effect or function of ‘particular lexical and grammatical choices’ which ‘generates expectations and inferences in communication and action’ (Semino et al. 2016: 3). In their study of representations of cancer on social media, Semino et al. (2016) build on the account of framing given in Conceptual Metaphor Theory (Lakoff and Johnson, 1980) as ‘a bias of conceptualisation’ which has the power to influence how we think about and act on a situation. While Semino et al. (2016) focus on how framing effects are achieved through metaphorical expressions, the definition they cite from Entman (1993) presents a broader perspective on this linguistic effect:

> Framing essentially involves selection and salience. To frame is to select some aspects of a perceived reality and make them more salient in a communicating text, in such a way as to promote a particular problem definition, causal interpretation, moral evaluation, and/or treatment recommendation for the item described. (Entman 1993: 52; italics in original)

In this article, we extend this notion of framing beyond metaphor to include non-metaphorical linguistic choices which contribute to how communication is construed more broadly in the health discourses we examine.

According to Semino et al. (2016: 11), analysis of framing should pay attention to not only entities, roles and relations being represented, but also ‘aspects such as agency, (dis)empowerment, evaluations, and emotional associations’. These framing implications are particularly significant in healthcare contexts (see also Baker et al. 2020; Demmen et al, 2015), where

> the choice of different descriptions of illness can have both positive and negative implications for the general well-being of people already in a vulnerable position. Here the emphasis is on the potential consequences for individuals of different (assumed) framings, and the goal is to develop guidelines or recommendations on what kinds of language should or should not be employed (Semino et al., 2016: 7)

This vulnerability is certainly true of those *experiencing*, who *have previously experienced*, or who *fear* pregnancy loss. The psychological and behavioural impacts of different ways of describing pregnancy loss using diagnostic terminology can be understood theoretically as framing effects (Malory 2025; Malory & Parr, 2025). In this article, rather than focusing on how the experience of pregnancy loss is framed, we focus on the framing of communicative interactions between patients and healthcare professionals about pregnancy loss. Our goal is to consider the potential ramifications of the framings under examination, for how practitioners and patients approach these communicative situations.

## 3. Data and methods

In order to investigate institutional discourse around pregnancy loss in the UK, two corpora were constructed. The first of these – the ‘Medical corpus’ – consisted of web content and online documents about pregnancy loss from 7 prominent UK medical organisations (see Appendix 1; Table 1). Since the research aims to identify possible implications of the language for public and practitioner conceptions of pregnancy loss care, texts were sampled on the basis of ease of accessibility and hence potential reach and impact.

**Table 1.** Breakdown of four sub-corpora by audience and wordcount.

| **Sub-corpus** | **Primary audience** | **Word count** |
|---|---|---|
| A: Medical- HP (healthcare-professional facing) | Healthcare professionals/trainees, commissioners and providers | 91,437 |
| B: Medical-P (patient-facing) | Patients, families, caregivers, and general public | 28,931 |
| C: Charity-HP (healthcare professional-facing) | Healthcare professionals/trainees, commissioners and providers | 12,162 |
| D: Charity-P (patient-facing) | Patients, families, caregivers, and general public | 212,158 |

Texts were sourced by searching for key terms in the websites of medical organisations such as the National Health Service (NHS), the National Institute for Health and Care Excellence (NICE) and the Royal Colleges of healthcare professions likely to be involved in pregnancy loss care. Key terms (*miscarriage, ectopic, stillbirth, pregnancy loss, baby loss, fetal loss, fetal death, termination)* were chosen on the basis of previous research into common terminology surrounding pregnancy loss in the UK (Malory, Nuttall and Heazell, 2025). Only publicly accessible texts were collected, meaning that any content requiring professional membership and/or password access were not included in the corpus.

Where a webpage or document covered broader topics such as maternity care or reproductive medicine, only sections relevant to the focus of the analysis were collected. This relevance was determined by the title and/or inclusion of two or more instances of at least one key search term in the body of the text. The same sampling criteria were applied to texts covering related topics, such as the abortion of unwanted pregnancies and neonatal death (the loss of a baby in the first 28 days after birth). The collected texts include patient information pages/leaflets, clinical guidelines, training materials/curricula for healthcare professionals, best practice papers, and reports from audits of clinical services. This range is representative of this discourse-type and the multiple registers typically contained with it, including research findings, (fictional) case studies, FAQs, resource lists and contact details. However, since our focus is on written text data, any videos or podcasts were not included.

In order to provide a point of comparison, a second corpus – the ‘Charity corpus’ – was built using similar methods. This corpus consisted of web content from 8 prominent UK charities on the subject of pregnancy loss (See Appendix 1; Table 2). The search terms listed above were used to collect publicly accessible written content on this topic, with the resulting texts including both patient information guides or booklets, and guidelines for healthcare professionals, and again featuring case studies and contact lists along with detailed information and advice. While this corpus does not contain the same range of text-types as the Medical corpus, this is again representative of this discourse, which is primarily focused on information and guidance for patients and their families.

**Table 2.** Key semantic domain comparison of the Medical corpus (target) with the BE21 (reference), sorted by significance (LL)

| **Semantic domain (USAS)** | **Frequency in Medical corpus:**<br>**(relative freq.)** | **Frequency in BE21 corpus:**<br>**(relative freq.)** | **Log Likelihood** | **LogRatio** |
|---|---|---|---|---|
| Q2.1 Speech: Communicative | 424 (0.29) | 8087 (0.66) | -334.76 | -1.16 |
| Q2.2. Speech acts | 1906 (1.33) | 10304 (0.84) | +300.96 | 0.66 |
| Q3. Language, Speech and Grammar | 156 (0.11) | 2042 (0.17) | -30.08 | -0.62 |

As Appendix Tables 1 and 2 show, our corpora do not contain equal amounts of text from each organisation sampled. The number of texts from each organisation varies, as does the length of these texts, reflective of the range of eligible content that was relevant to our topic from each source, as defined by our sampling criteria. The analysis does not make claims about specific organisations, but rather the dataset as a whole, and findings must be interpreted in relation to the make-up of each corpus. Finally, these samples are not exhaustive of the institutional discourse on pregnancy loss published online. However, since our aim was to collect texts most likely to reach the widest audience, sampling prioritised accurate representation of the most prominent, or influential organisations.

The two resultant corpora contain 120,383 words (Medical corpus) and 224,481 words (Charity corpus), respectively. While relatively small, these datasets are intended to be representative of these two types of health discourse for the purposes of corpus analysis. However, the different audiences targeted within each corpus (see Appendix 1; Tables 1 and 2) poses challenges for comparison. The role of third sector organisations means that the Charity corpus is dominated by texts aimed primarily at patients, families and the general public, with only a smaller number targeting healthcare professionals specifically. By contrast, the Medical corpus is dominated by texts aimed at healthcare professionals, trainees, or commissioners, with a smaller number targeting patients and the general public. To address this challenge, both corpora were divided into two sub-corpora, with texts categorised based on their function of information sharing: ‘healthcare professional-facing’ or ‘patient-facing’. Word counts for the four sub-corpora are shown in Table 1.

All corpora were cleaned and converted to .txt files suitable for analysis using Wmatrix7 corpus analysis software (Rayson, 2009). Frequency, semantic keyness, keyword and concordance analyses were conducted using Wmatrix7 in multiple phases of analysis.

Phase 1 focused on the representation of communication in the Medical corpus. After an initial examination of word frequencies for specific terms, the Medical corpus was tagged using Wmatrix7’s USAS Semantic Tagger, which automatically assigns the semantic domains expressed by words based on a pre-existing lexicon (Archer et al., 2002). This was then compared to the semantic domains tagged in a reference corpus, ‘BE21’: a sample of 1 million words of general written British English published in 2021 (Baker, 2023). Those semantic domains that were significantly more or less frequent in the Medical corpus relative to the BE21 reference corpus (those that are ‘key’, Bondi and Scott, 2010) were then manually examined to identify any domains relating to communication.

In Phase 2, the Medical corpus and Charity corpus were compared. A list of keywords, or words which exemplify the ‘aboutness’ of the texts contained (Stubbs, 2010), were identified for each corpus. ‘Positive’ keywords in each corpus (those words that are statistically more frequent) were calculated relative to the other as reference corpus, with a minimum Log Likelihood of 6.63 (p<0.01), a minimum LogRatio of 1^1^ and a minimum frequency of 1. Results were sorted by effect size (LogRatio) and the top 150 results in terms of effect size sampled.

In Phases 3 and 4, keyword analyses were again conducted, this time comparing the pairs of sub-corpora matched in terms of target audience (Table 1). Owing to the small size of the Charity-HP and Medical-P sub-corpora (see Table 1), these were each selected as the target corpus in their respective comparisons. Both positive and negative keywords (those that are significantly more and less frequent in the target corpus, respectively) were identified relative to the larger audience-matched reference corpus. Keywords were collected, sorted and sampled using the same procedures as Phase 2^2^. For these comparisons, the small size of the sub-corpora means that caution must be exercised in the interpretation of statistical significance. Here, effect size (based on relative frequency of use) offers a useful measure of linguistic difference.

Finally, concordance lines for each (sub)corpus were manually examined for those positive and negative keywords relating to communication, with a focus on the specific ways in which interactions between patients and healthcare professionals are framed in context.

## 4. Analysis

These corpus linguistic methods allowed us to assess the extent to which communication is a topic within the institutional health discourses, and the way in which it is framed when it does appear. Results from each phase of analysis are outlined in the following four sections.

### 4.1 Communication without dialogue?

In order to assess the extent to which the topics of language and communication are discussed in the Medical corpus (RQ1), a basic word frequency analysis was conducted using Wmatrix7. The lemmas *communic*\* and *lang*\* are seen relatively infrequently in the 120,383-word Medical corpus, with words such as *communicate, communicating, communication* appearing 32 times and mentions of *language(s)* appearing just 17 times. This may reflect the under-privileging of communication as a factor in perinatal healthcare (see Malory, 2025).

However, references to these topics can come in many other forms and raw frequencies do not tell us whether this usage is high or low relative to general British English. In order to explore how often the *concept* of communication is discussed in the data, a key semantic domain analysis was conducted (see Section 3). As we might expect, domains such as ‘Anatomy and physiology’, ‘Medicines and medical treatment’ and ‘Disease’ were those most significantly overrepresented, or most key, in the Medical corpus compared to general written British English; while ‘Pronouns’ and ‘Personal names’ were those most significantly underrepresented. Results for those semantic domains most relevant to the topic of language/communication are given in Table 2.

This preliminary analysis shows that the semantic domain of ‘Speech acts’, definable as an utterance that serves a communicative function, occurs significantly *more* frequently in the medical corpus relative to general British English. Examination of concordance lines for this domain shows words such as *recommendation(s)*, *advice, explain, report* and *referral* tagged in this category. On the other hand, both the domains of ‘Speech: Communicative’ (containing ‘terms relating to spoken communication’ (Archer et al, 2002: 23); e.g. *talk, speak, communication*) and ‘Language, Speech and Grammar’ (containing ‘terms relating to language’ (2002: 24); e.g. *language, words, terms, read, interpreter*) both occur significantly less frequently in the Medical corpus relative to general written British English. While communication does appear, therefore, to be a topic within the medical discourse, this analysis suggests that certain kinds of communication (particularly those kinds carried out by professionals) are emphasised over others, and that the speech and language through which communication is achieved may not be a primary focus. The *type* and *nature* of the communication described in this corpus, or the ways in which the different interactions which take place in this context are ‘framed’ (see Section 2), are therefore in need of closer analysis.

### 4.2 Communication in the discourse of medical organisations vs charities

In order to compare how communication is framed (RQ2), the Medical Corpus was then compared to the 224,481-word Charity corpus, this time looking for positive keywords in order to examine the specific expressions at work in construing communication in each discourse (see Section 3). Sampled keywords for each corpus were manually grouped into categories based on common themes (see full samples in Appendix 2, Tables 1 and 2).

Very few communication-related keywords appear in the Medical corpus (Table 3), which is instead predominately characterised by words reflecting clinical terminology, scientific data and formal policy. While *recommendation* and *recommendations* appear as key, these are most often used in relation to policy as opposed to clinical interaction. In the Charity corpus, on the other hand, words relating to various types of spoken and written communication are overrepresented relative to the Medical corpus, alongside words referring to emotional responses/mental states and memorialisation after loss.

**Table 3.** Positive keywords in the Medical corpus.

| Category | Example keywords |
|---|---|
| **Clinical terminology for conditions, medication or treatment** | *analgesia, biochemical, dichorionic, endometrial, intrauterine, GTD, monochorionic, obesity, perinatal, preterm, resection, serum, tract, trophoblast* |
| **Scientific data and evidence** | *births, compared, concluded, controls, data, evaluation, evidence, figure, interpretation, proportion, population, rates, studies, table, trial* (also dates and percentages) |
| **Policy and formal guidelines** | *agreed, amended, categories, classification, committee, consensus, guideline, practice, rationale, recommendation(s), strength* |
| **Pregnancy outcomes** | *abortions, adverse, morbidity, mortality, outcomes, survival, terminated* |
| K**nowledge (professional)** | *assessing, accuracy, detection, determined, expertise, indications, suspicion* |
| **Graded measures** | *severity, status* |
| **Professional roles** | *practitioners* |

The words *say* and *talking* are two of the top three most statistically overrepresented keywords in the Charity corpus (*say*: p<0.0001; LR 6.2, *talking*: p<0.0001; LR 5.1; see Appendix 2, Table 2). *Talking* appears only twice in the Medical corpus, in both cases in the context of ‘talking to’ women and families:

**Figure 1.**
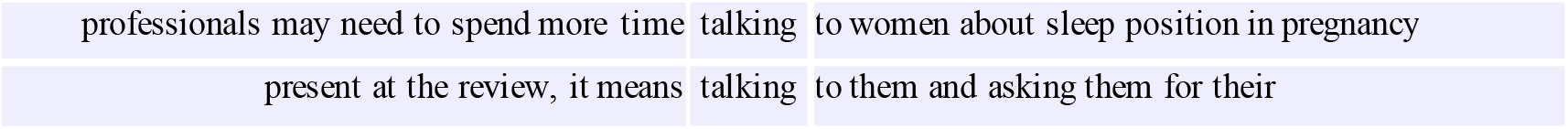
All concordance lines for *talking* from the Medical corpus (total 2 instances).

Contrastingly, in the Charity corpus, the act of *talking* is represented in a variety of ways across 132 instances. Here, the person experiencing loss, or a member of their immediate family, is most often the speaker (in 110 out of 132 instances), talking ‘to’ healthcare professionals, family members and colleagues, ‘about’ a range of specific topics relating to pregnancy loss. Figure 2 exemplifies this, with a sample of 10 concordance lines from the Charity corpus.

**Figure 2.**
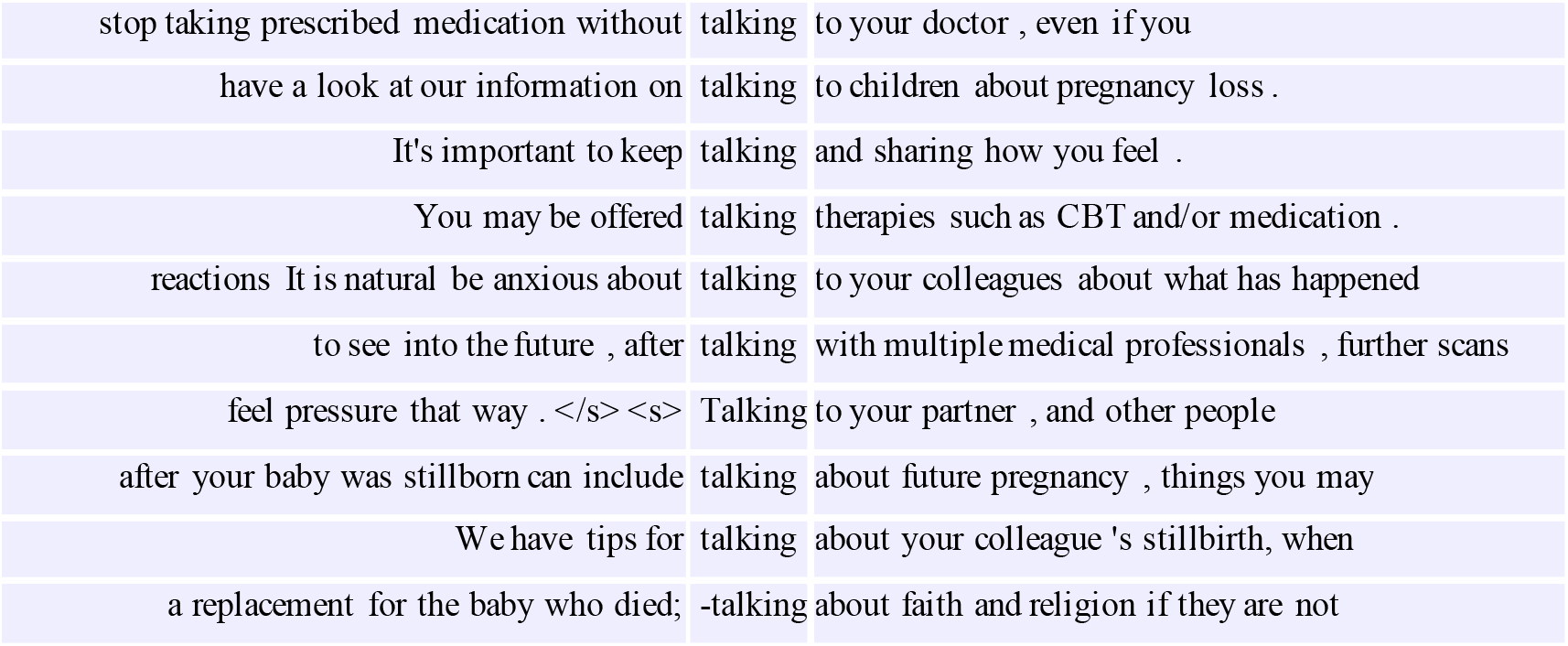
Sample concordance lines for *talking* from the Charity corpus (total 132 instances).

Alongside spoken communication and its reception (*listen, hear*), forms of written communication received by patients such as *instructions* on how to take medicines, a *letter* received from hospitals or practitioners, and opportunities to submit *feedback*, appear as keywords in the Charity corpus (Table 4).

**Table 4.** Positive keywords in the Charity corpus.

| **Category** | **Example keywords** |
|---|---|
| **Emotions and mental states** | *angry, comfortable, cope, emotions, feelings, frighten, frightening, gentle, hard, heal, helped, kind, knew, knowing, sadness, thoughts, worried, worries* |
| **Memorialisation** | *ashes, ceremony, crematorium, funeral, gone, grave, memories, picture, remember* |
| **Spoken and written communication** | *Feedback, instructions, hear, letter, listen, reassurance, share, sharing, say, says, talk, talking, tell, telling, told* |
| **Professional roles** | *colleague, counsellor, director, employer, sonographer* |
| **Pronouns** | *his, him, me, our, us, yourself* |

The final two categories of keywords in the Charity corpus: ‘professional roles’, and ‘pronouns’, are also relevant to communication. The former category reflects mention of specific interactions with professionals occurring along the care pathway, often in the context of explaining their role to the recipient of care (e.g. *sonographer*; p<0.001; LR 2.98, examples in Figure 3).

**Figure 3.**
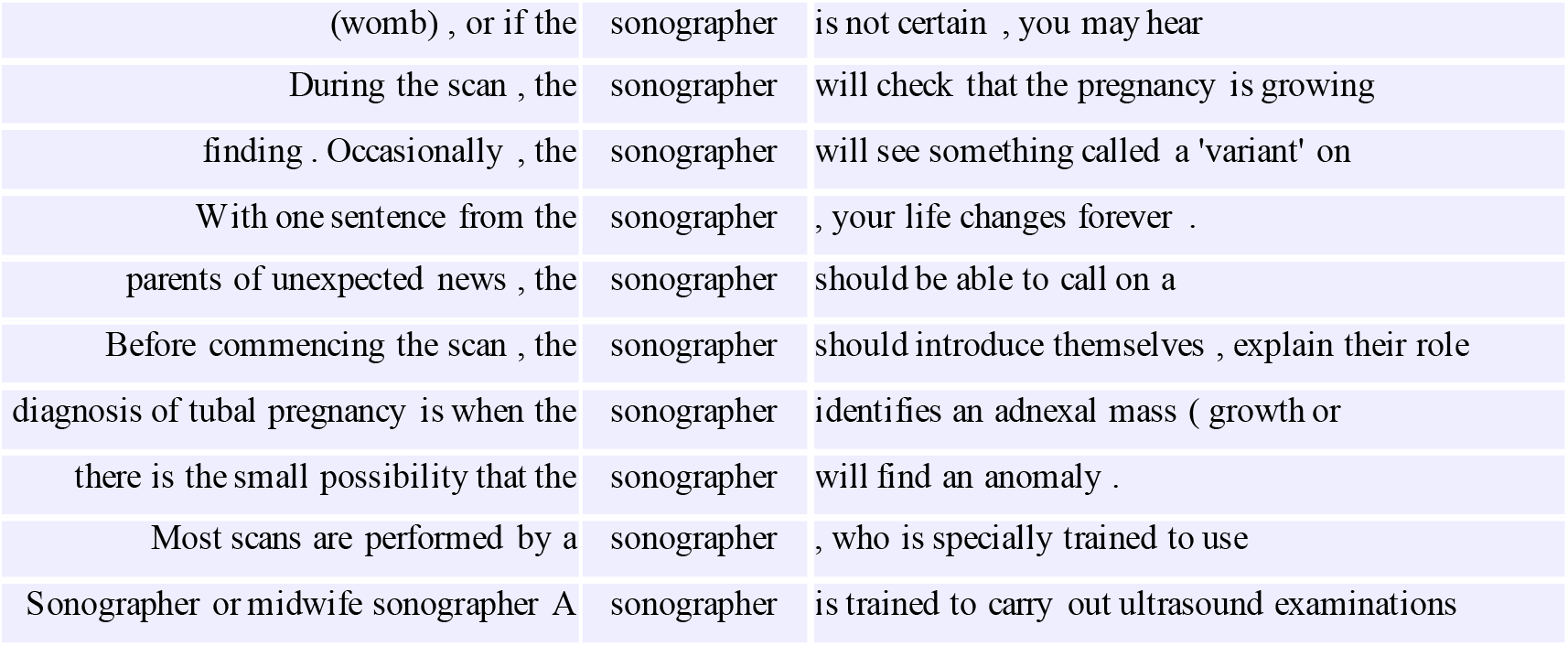
Sample concordance lines for *sonographer* from the Charity corpus (total 30 instances).

In the Medical corpus, the collective term *practitioners* instead occurs as a positive keyword (p<0.0001; LR 4.18), in contexts where requirements or expectations of staff are set out in broader terms. References to specific professional roles such as *sonographer* are rarely found (and do not appear as keywords). For example, just 2 instances of *sonographer* are seen, only one of which describes an interaction between professional and patient:

**Figure 4.**
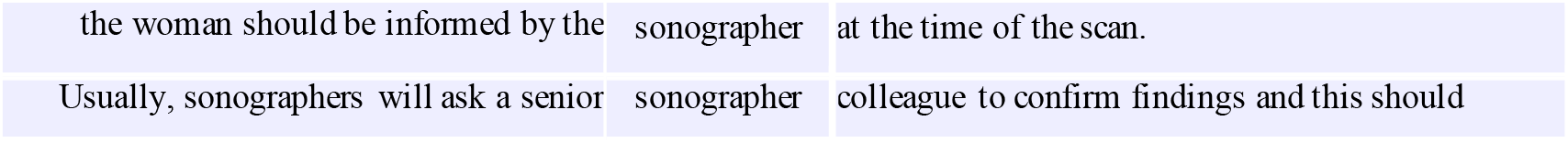
All concordance lines for *sonographer* from the Medical corpus (total 2 instances).

The final keyword category (‘pronouns’) often relates to the voice of the parents or families themselves, in written testimonies or podcasts referenced within the corpus, and sometimes reflects their own way of referring to the baby they have lost (e.g. ‘we held his body, made decisions about his body, and organised our son’s funeral’ (Charity corpus).

Comparison of these two corpora has therefore clarified the findings of the preceding analysis in section 4.1. Numerous communication-related keywords are over-represented in the Charity corpus, which highlight different forms of professional-patient communication. That these appear significantly less frequently in the Medical corpus supports the finding that the topic of communication is given less attention overall within this discourse.

However, based only on these results, we cannot conclude whether the Medical and Charity corpora keywords, and the different degree of emphasis they place upon communication, reflect meaningful differences between the two text-types. It is possible that the patterns of keyness identified may instead be due to differences in the functions and audiences of the texts sampled (see Appendix 1). To explore further, the next phase of analysis focused on comparison of the two healthcare professional-facing sub-corpora and the two public-facing sub-corpora, separately (Section 3, Table 1). By comparing sub-corpora with similar audiences, and hence similar functions, we aim to identify patterns in how communication is framed for healthcare professionals and patients.

### 4.3 Communication in healthcare professional-facing texts

In order to examine how communication in the context of pregnancy loss care is framed for a healthcare professional audience, a closer comparison of the two healthcare professional-facing sub-corpora: A (Medical-HP) and C (Charity-HP) was then carried out (see Section 3). Positive and negative keywords sampled from the Charity-HP target corpus relative to the larger Medical-HP corpus were manually grouped into categories (see full samples in Appendix 2; Tables 3 and 4).

The positive keywords in the Charity-HP corpus (Table 5) again reflect the prominence of themes relating to communication. Keywords relating specifically to formal communication received by patients, such as a *certificate, letter* or *postmortem* and other sources of private and public health information in *notes* and *leaflets,* distinguish this discourse from the Medical-HP sub-corpus, where mention of these documents, in which loss is described and *documented,* are significantly less frequent. The *wording* of such communication is also a positive keyword in the Charity-HP corpus (p<0.01: LR 3.33), albeit appearing in just 4 instances (Figure 5):

**Figure 5.**
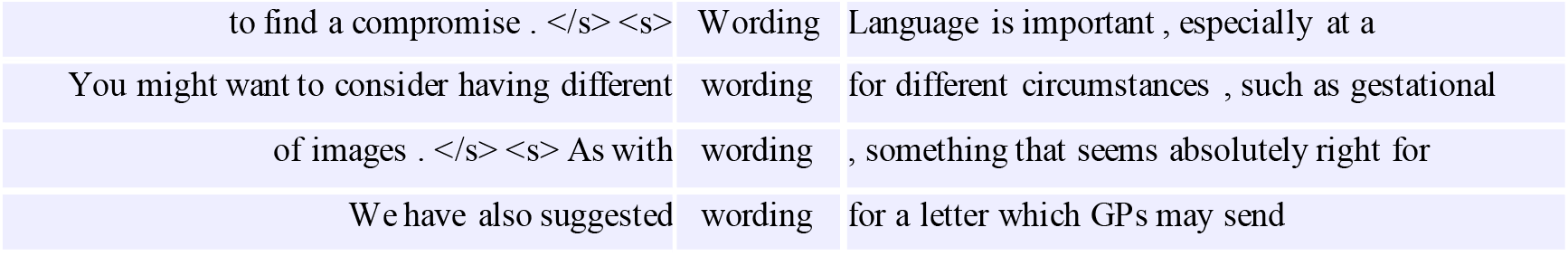
All concordance lines for *wording* from the Charity-HP corpus (total 4 instances).

**Table 5.** Positive keywords in Charity-HP sub-corpus.

| **Category** | **Example keywords** |
|---|---|
| **Communication – written/formal** | *certificates, certification, documented, leaflets, letter, notes,* (post) *mortem, publications, questionnaire, read, template, wording,* |
| **Communication – general** | *agree, call, communicating, confirmation, consultation, discuss, news, sensitively,* |
| **Professional roles and organisations** | *hospitals, involvement* (of other teams/services), *NHS, private* (providers), *team, teams, sector, sonographers, specialised* |
| **Laws (affecting healthcare professionals)** | *right, rights, legally,* (NHS) *contract* |
| **Individual preferences and feelings** | *appreciate, feel, helpful, individualised, memory, reassured parents, prefer, want* |
| **Knowledge (patient)** | *prepare, know, uncertainties, expect, unexpected* |
| **Knowledge (professional)** | *assumptions,* (training) *sessions, think* |

Although the number of instances for this keyword is small across both sub-corpora, the 3 instances of this word in the larger Medical-HP corpus all relate to the updated *wording* of published guidance for healthcare professionals, as opposed to the language used in documentation for patients, indicating that guidance on formal communication methods for practitioners is underrepresented, and perhaps absent from, these recommendations.

Other keywords relating to communication more generally, such as *news*, *communicating*, *discuss,* are used significantly more often in the discourse aimed at healthcare professionals by charities compared to that produced by medical organisations. Unlike the largely patient-focused keywords such as *talking, say* and *tell* which characterised the Charity corpus as a whole (see Section 4.2), most instances of these keywords relate to the manner in which information is communicated by the healthcare professional to the patient, with the professional represented agentively, as speaker. For example, ‘*Communicating* unexpected or difficult *news* is challenging and all sonographers should have..’(Charity-HP corpus); ‘Fetal Medicine doctor to *discuss:* further imaging e.g. ultrasound surveillance..’ (Charity-HP). As an exception to this, *discuss* is used in a minority of cases in both sub-corpora to frame the patient as communicator: ‘some women may want to *discuss* their decision further with their local obstetrician’ (Medical-HP); ‘may be helpful for parents who wish to *discuss* termination with someone’ (Charities-HP), or to frame both patient and professional as implicit communicative participants: ‘A face-to-face appointment to *discuss* ongoing care should be offered as soon’ (Charity-HP); ‘an appointment to *discuss* postmortem results needs to be arranged’ (Medical-HP). In these interactions, agency appears more evenly distributed between patient and professional, framed as a two-way exchange of information, feedback and decision making. However, these opportunities for discussion are typically authorised (‘offered’ or ‘arranged’) by the healthcare professional in a preceding communicative act.

The keyword *sensitively* (also *sensitive*) is used to describe how such communication with patients should be carried out. Although once again appearing in small numbers in both sub-corpora, this keyword is overrepresented in the Charity-HP sub-corpus relative to the Medical-HP sub-corpus (p<0.001; LR 2.72):

**Figure 6.**
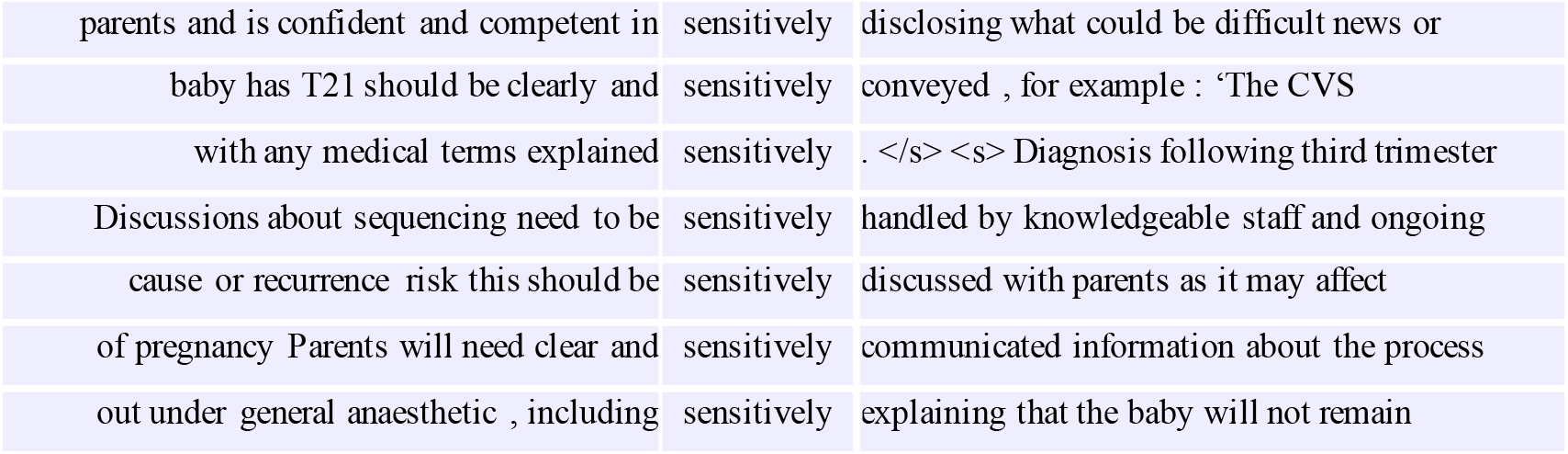
All concordance lines for *sensitively* from the Charity-HP corpus (total 7 instances).

Other keywords relating to the ‘Individual preferences and feelings’ of patients (see Table 5) – including the keyword *parents* alongside *appreciate, feel, want, prefer –*contribute in similar ways to the discussion of communication within the Charity-HP sub-corpus. This framing of the healthcare professional as agentive communicator, in the evaluative context of what ‘should’ be communicated and how this ‘needs to’ be done, is in line with the function of the texts in this sub-corpus, which set out information and recommendations for healthcare professionals on different aspects of pregnancy loss care. Despite sharing this same function, the Medical-HP sub-corpus places significantly less emphasis upon these particular aspects of the healthcare professional role.

This analysis supports the interpretation that spoken and written communication of various kinds are given greater attention in the discourse on pregnancy loss from charities, than that from medical organisations. By comparing the portions of the corpora aimed at healthcare professionals only, we have been able to rule out the possibility that keyword differences are related to functional differences in the texts sampled. Based on these findings, we can argue that while communication has been shown to be a vital component of care in maternity contexts (see Section 1), existing institutional discourse by UK medical organisations aimed at healthcare professionals in this setting does not currently reflect this stance. In contrast to health discourse on pregnancy loss from charities, which contains clear, evaluative guidance on *how* professionals should communicate with those who experience pregnancy loss, both orally and in writing, medical discourse appears to offer very limited guidance for professionals. The possible implications of this discourse framing for the professional identity of healthcare professionals, along with their attitudes and priorities in practice, will be returned to in Section 5.

### 4.4 Communication in patient-facing texts

A parallel comparison of the two patient-facing sub-corpora: B (Medical-P) and D (Charity-P) was carried out in order to examine how communication in the context of pregnancy loss care is framed for patients, families and members of the general public seeking advice and information. Positive and negative keywords sampled from the Medical-P corpus relative to the larger Charity-P sub-corpus (see Section 3) are again grouped into categories below (full keyword samples in Appendix 2, Tables 5 and 6).

**Table 6.** Negative keywords in Charity-HP sub-corpus.

| **Category** | **Example keywords** |
|---|---|
| **Terminology for specifying types of loss** | *cervical, early, ectopic, gestational, late, maternal, miscarriage, pregnancy/cies, preterm, recurrent, stillbirth, uterine* |
| **Policy and research** | *evidence, rationale, recommendations, research, review, studies* |
| **Medical procedures** | *assessment, caesarean, examination, management, treatment, ultrasound* |
| **Gendered language** | *she, woman, women* |

Unlike the healthcare-professional facing sub-corpora analysed in the previous section, communication is a shared topic across both public-facing sub-corpora, since keywords relating to ‘Communication’ appear in both Tables 7 and 8. However, the two sets of keywords differ in how they frame patient-professional interactions. Echoing the findings of the key semantic domain analysis in section 4.1, certain types of communicative situations are represented in the Medical-P discourse while others are underrepresented or missing. Specifically, words relating to a one-way transfer of information from the healthcare professional to the patient: e.g. *advise(d), recommended, offered,* are significantly more frequent in the Medical-P corpus. Meanwhile, words which express two-way communication between the patient and healthcare professional, e.g. *talk, tell, say, ask,* are significantly less frequent relative to the Charity-P corpus. Examination of concordance lines illustrates the framing that results from the former group of linguistic choices:

**Table 7.** Positive keywords in the Medical-P sub-corpus.

| **Category** | **Example keywords** |
|---|---|
| **Risk and lifestyle choices** | *alcohol, avoid, bacterial, contraception, diet, drinking, drugs, flu, food, increase* (risk), *obese, overweight, poultry, raw, reduce* (risk), *smoking, tampons, vegetables* |
| **Communication** | *advice, advised, advisable, answering, confirm, confirmed, discuss, follow-up, known* (as), *offered, recommended, reporting, suspected,* (medical) *terms* |
| **Clinical terminology for conditions, medication or treatment** | *cholestasis, foetal, gtd, laparoscopy, pcos, pre-eclampsia thrombophilia, trisomy* |
| **Cause and prevention (of miscarriage)** | *cause*, *harm* (baby), (baby’s) *movements, preventing, reporting, sign* |
| **Gendered language/gender identity** | *gender, identity, inclusive, mother, ‘woman’, ‘women’* |
| **Body of the birthing parent** | *fallopian, hands, tubes, tummy, uterus, vaginal, weakened (cervix)* |
| **Emotions and mental states** | *Affects, recovering,* (come to) *terms,* |
| **References to the baby** | *foetal, foetus, non-tubal (ectopic pregnancy), tissue* |
| **Patient choices** | *choices, options* |

**Table 8.** Negative keywords in the Medical-P sub-corpus.

| **Category** | **Example keywords** |
|---|---|
| **Emotions and mental states** | *upset, upsetting, difficult, children, caring, cope, feel, feelings, emotions, experiences, hard, grief, loss, memory, mental* |
| **Pronouns** | *their, them, they, our, we, me, my, I, yourself, her* |
| **Communication** | *ask, describe, hear, say, share, talk, tell,* |
| **Gender neutral language** | *everyone, partners, people, parents, person* |
| **Knowledge (patient)** | *understanding, know, remember* |
| **Formal processes** | *post-mortem, report, right* |
| **References to the baby** | *babies, fetal* |
| **Medical professionals** | *doctors, staff* |

The examples in Figure 7 reflect patterns in the use of these keywords, which is typical of the wider Medical-P sub-corpus. The healthcare professional is typically the agentive communicator in each process, responsible for providing advice or recommendations, though the grammatical structures in which these words are used often background this professional participant. This includes the use of the passive voice, as in ‘you will be advised’ or ‘you may be offered’, where it is the passive patient-recipient ‘you’ who is foregrounded in subject position, with the professional who advises or offers scans, grammatically absent from the clause altogether. In other instances, both patient and professional are removed from attention: for example, ‘methotrexate may be *recommended*’, ‘it is *advisable*’, ‘miscarriage is *confirmed*’ do not attribute the implicit communicative (and interpretative) processes described by these verbs to any particular participant, and in some cases attribute this process to a ‘test’ or ‘results’ as opposed to a person who interprets and delivers information: e.g. ‘results can confirm’. This is also the case for nouns such as *advice* and *follow-up*, in which an interaction between patient and professional is implied but not made salient by the framing. While the majority of communicative keywords characteristic of the texts produced by medical organisations for public audiences therefore again frame healthcare professionals in an agentive role in interaction with patients, this analysis raises a further consideration here concerning the explicitness with which this professional communicator is construed within patient-facing information. Alongside the attribution of *agency*, the *saliency* with which (specific) healthcare professionals are represented as communicators at particular points in care has other implications for the patients who are recipients of such texts.

**Figure 7.**
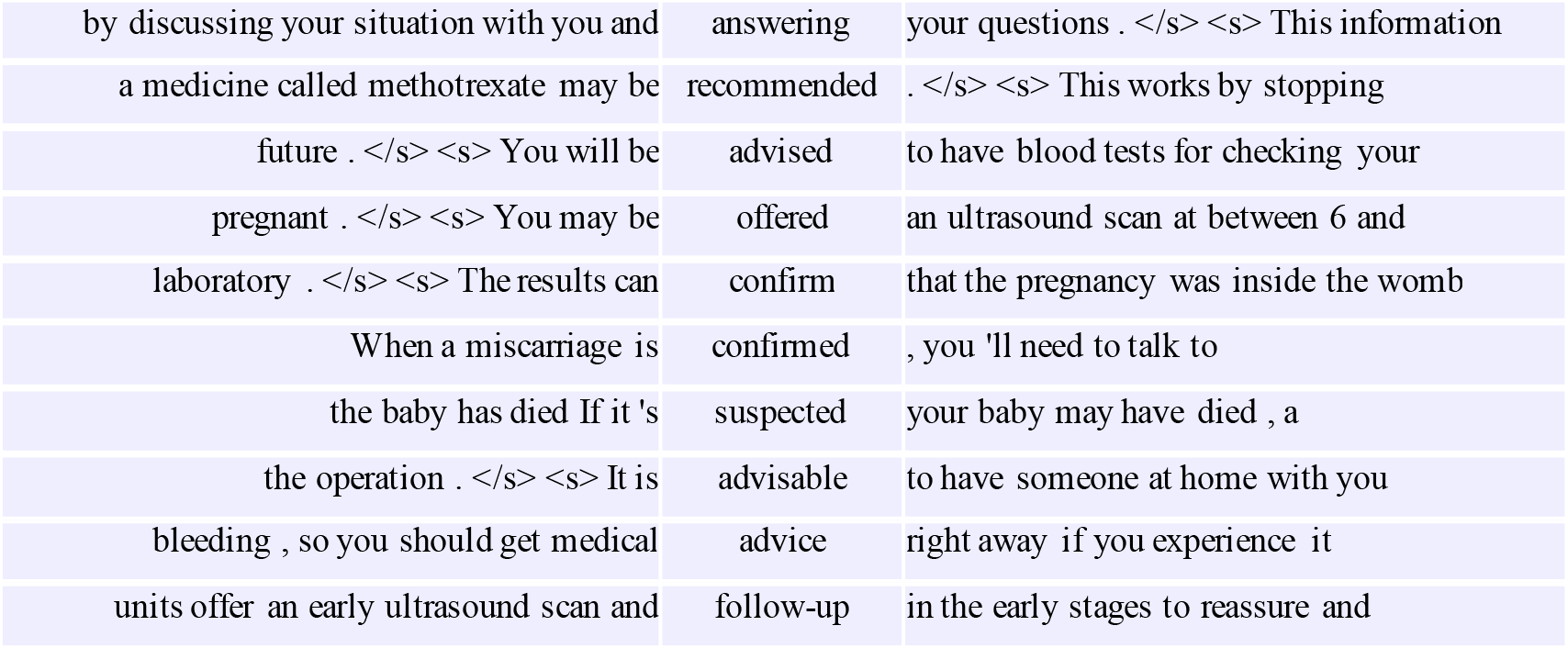
Sample concordance lines for communicative processes from the Medical-P corpus.

One notable exception to this framing pattern is *reporting.* This keyword is overrepresented in the Medical-P corpus (p<0.0001; LR 4.87), where it is consistently used to frame the patient both as communicator and as responsible for surveillance of their pregnancy symptoms, in the context of reporting concerns:

**Figure 8.**
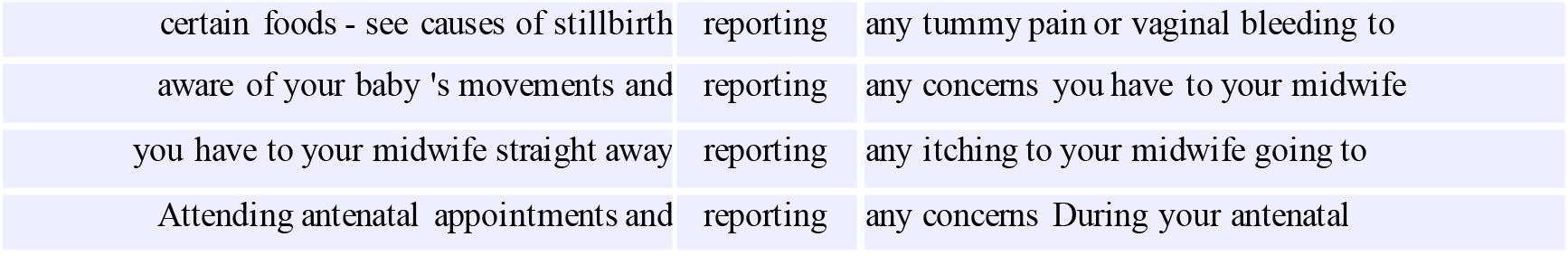
All concordance lines for *reporting* from the Medical-P corpus (total 4 instances).

Emphasis on this communicative act can be interpreted in light of the keyword categories: ‘Risk and lifestyle choices’ and ‘Cause and prevention’, which characterise the Medical-P sub-corpus (Table 7), as well as broader societal discourses of an individual’s responsibility for ensuring their pregnancy progresses safely (Malory, 2027). The discourse aimed at patients and the general public produced by medical organisations appears to place a much greater emphasis upon knowledge of, and communication in response to, risk factors for pregnancy loss, than the comparable discourse produced by charities. Since this is one of very few communication-related keywords in the Medical-P sub-corpus that frames the patient as agentive communicator (also some instances of *discuss*, use of which is comparable to that seen in Section 4.3) – and the only one to do so exclusively – this finding highlights the possibility that only specific kinds of communication, and hence only specific kinds of power or influence, are being attributed to patients in relation to their pregnancies and their care, as one implication of this discourse framing.

Looking next to those negative keywords that are used significantly less frequently in the Medical-P sub-corpus (Table 8), we see a range of verbs which describe (spoken) communication: e.g. *talk, tell, say.* The way in which communication is framed by these word choices is illustrated by the concordance lines below:

These verbs describe communication in both directions between the patient and healthcare professionals involved in their care, as well as between patients and their families, friends and colleagues. These word choices are less formal compared to the communication-related keywords which characterise the Medical-P corpus previously examined (see Figure 9). Notably, they are more active; simple (e.g. ‘nurse might *ask*’) and infinitival (‘to *say’*) verb forms, occurring in the active as opposed to passive voice (e.g. ‘the hospital staff will *tell* you’, rather than ‘you will be told’). Through this framing, communicative agency is assigned to <u>both</u> patients and professionals using the same linguistic choices, with the specific participants involved in the interaction made salient in attention.

**Figure 9.**
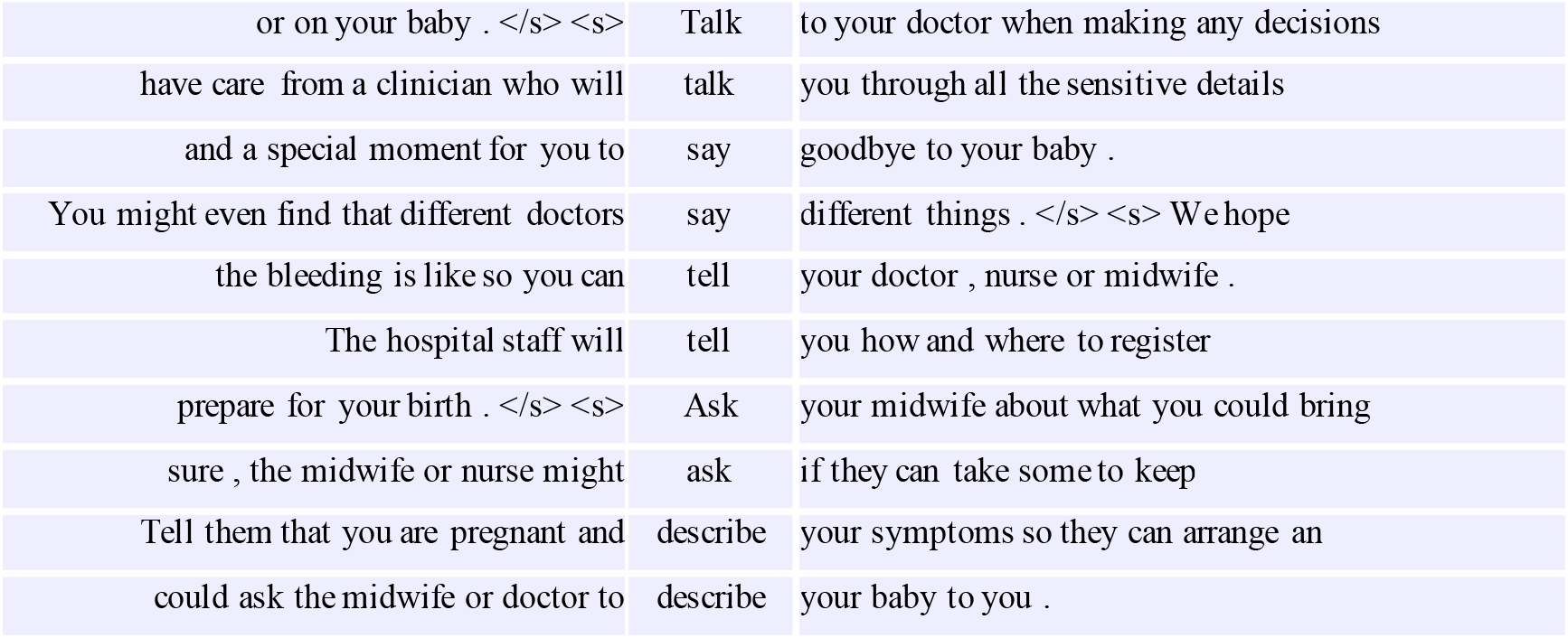
Sample concordance lines for communicative processes from the Charity-P corpus.

Framing of interaction therefore seems to exhibit key differences between the two patient-facing sub-corpora. Firstly, patient-facing texts produced by charities more frequently frame the patient as an agentive communicator who shares, questions or describes their experiences; often through the same communicative processes attributed to the healthcare professional (e.g. *talk, tell, say*). Patient-facing texts produced by medical organisations, on the other hand, typically attribute agency or power to the professional, except for in specific scenarios such as the *reporting* of risk. This provides evidence that charities’ discourses are more empowering in their more balanced framing of communication as reciprocal and bidirectional.

Secondly, medical organisations’ outputs focus primarily on conveying a diagnosis, treatment or advice, with the professional often backgrounded, or implicit, as the communicator in a transfer of information to the patient (e.g. ‘the results *confirm*’). By contrast, the charities’ outputs regularly foreground individual healthcare professionals (e.g. *sonographers*, Section 4.2) as active communicators in specific interactions with patients (e.g. ‘a clinician will *talk* you through’). In this way, patient-facing texts produced by charities can be seen as more transparent and accessible in their representations of typical interactions and what can be expected from professionals involved in care.

Comparing these two patient-facing sub-corpora has therefore revealed differences in both the types of communication being represented and the extent to which participants, as active communicators and respondents, are foregrounded through the language used. This could have ramifications for patients and other members of the public who access these texts to better understand the experience of loss.

## 5. Conclusions

This study examined the extent to which communication is discussed in UK institutional discourses around pregnancy loss (RQ1), and how communication between patients and healthcare professionals is framed within these discourses (RQ2). The findings reveal a consistent contrast between texts produced by medical organisations and those produced by charities. In the Medical corpus, semantic domains relating to spoken communication and language were significantly under-represented relative to general written British English. Although speech acts were over-represented, these primarily concerned institutional or unidirectional professional communication modes, such as advising, recommending, and referring. Communication was therefore predominantly construed as the transmission of professionally authorised information rather than as dialogue. The keyword comparison reinforced this finding. Words relating to dialogue between patient and professional (e.g. *say, telling, hear, listen)*, were over-represented in the Charity corpus along with mentions of specific professional roles involved in key conversations. Overall, charity outputs gave greater prominence to talking, listening, feelings, preferences, wording and patient perceptions. Medical outputs instead foregrounded clinical terminology, evidence, policy, risk and professional knowledge.

The audience-matched keyword analyses clarified how this contrast manifested in texts aimed at different audiences. Across healthcare professional-facing texts, those produced by charities again featured a greater emphasis on forms of written and spoken communication as part of clinical practice, including discussion of wording or delivery, and the needs and preferences of parents. As well as giving less emphasis to communicative aspects of care, texts produced by medical organisations also featured a lack of guidance as to how they ought to be carried out by professionals. Across patient-facing texts, key differences between medical and charity discourses related to the framing of specific types of communication. Charity outputs represented both patients and professionals as communicatively active and more frequently identified (likely) individual participants in particular interactions. In Medical outputs, characteristic communicative expressions, such as *confirmed, advised, recommended* and *offered*, foregrounded professional agency in these communicative scenarios, but typically did not identify the individual participants involved. Patient agency was most clearly foregrounded in relation to *reporting* symptoms or concerns. Taken together, these patterns frame medical communication as a largely unidirectional, and often impersonal, transfer of information, while charity discourse more often frames it as interpersonal, reciprocal, and emotionally consequential.

Several limitations qualify these conclusions. The corpora are relatively small, unequal in size and restricted to publicly accessible written materials; they therefore exclude password-protected professional resources and spoken or multimodal content. Corpus patterns cannot demonstrate how individual texts are received or establish direct effects on clinical practice. Future research should examine whether these framings recur in training materials and clinical interactions, and should test their effects through interviews, observations and co-produced evaluations with healthcare professionals and people with lived experience.

### 5.1. Implications for healthcare professionals and professional bodies

Institutional discourse contributes to defining what healthcare professionals are expected to know and do. The Medical corpus constructs the professional’s communicative role primarily as the transmission of clinical knowledge: someone who diagnoses, advises, recommends and offers treatment. Far less attention is given to the professional as a relational communicator who listens, responds to emotion, explains terminology sensitively, elicits preferences, and checks understanding. Communication is consequently positioned as peripheral to clinical expertise rather than as one of the means through which safe and compassionate care is delivered.

The recurrent use of passive and agentless constructions compounds this problem. Expressions such as “you will be advised” or “miscarriage is confirmed” obscure who communicates information, how it is communicated, and who is responsible for ensuring that it has been understood. They can also represent diagnoses and recommendations as impersonal clinical facts rather than information delivered by particular professionals to particular people in circumstances of vulnerability. This framing does not mean that healthcare professionals themselves communicate in these ways, but it means that the institutional texts intended to guide them provide limited discursive support for recognising communication as a core component of their professional role.

Medical organisations should therefore integrate specific communication guidance throughout clinical guidelines, curricula, and patient pathways; rather than treating it as a separate or optional “soft skill”. Guidance should identify who is responsible for communicating information, specify the importance of listening and responding as well as explaining, address the emotional and interpersonal effects of wording; and require professionals to elicit language preferences, invite questions, and check understanding. The greater attention paid to these practices in charity-produced professional guidance demonstrates awareness within that sector of the need for this guidance. Charities should not, however, be left to compensate for omissions in authoritative medical discourse: responsibility for establishing communication as a clinical standard rests with the institutions that define and regulate professional practice.

### 5.2. Implications for patients

The framing identified in patient-facing medical discourses also has implications for what patients understand themselves to be entitled and expected to do. When communication is represented principally through being *advised, offered* or *told*, patients are positioned primarily as recipients of professional knowledge. Their communicative agency is most visible when they are instructed to report symptoms or risk indicators. This creates a restricted model of participation in which the patient’s role is to supply clinically relevant information and receive professional decisions, rather than to question, interpret, express preferences, or contribute experiential knowledge.

The prominence of risk and lifestyle discourse may further individualise responsibility for preventing adverse outcomes, while the backgrounding of professional communicators makes institutional responsibility less visible. By contrast, charity discourses provide patients with a more explicit account of what interactions may involve, who will communicate with them and what they may wish to ask, say, share, or know.

Patient-facing information from medical organisations should therefore represent communication as an active, two-way process. It should name the professionals involved, explain what patients can expect from them and legitimise questions and preferences. Such changes would not simply make information more compassionate or accessible. They would challenge a paternalistic framing in which professional knowledge is privileged and patient knowledge is marginalised. If listening to women and birthing people is to be treated as a critical safety issue, this principle must be visible in the institutional language through which pregnancy loss care is described, taught, and enacted.

## Ethics approval statement

This article relies entirely on existing materials in the public domain and does not involve any experiments, interventions, or data collection involving human or animal subjects. Consequently, formal ethical approval was not required.

## Data availability

The data used in this research is in the public domain and accessible (at the time of submission) using the links provided in Appendix 1.

## Footnotes

1 A LogRatio (effect size) score of >1 means that the word appears at least twice as often in the target corpus compared to the reference corpus.

2 In the case of negative keywords, a maximum Log Ratio of -1 was the criteria for sampling. This meant that only those words that appeared at least twice as often in the reference corpus, compared to the target corpus, were sampled. In both comparisons, this resulted in a sample of less than 150 negative keywords (see Appendix, Tables 4 and 6.

