## Supplemental material: Appendix 1 for "Communication without dialogue? Framing patient–professional interaction in UK institutional discourse on pregnancy loss"

| **Table 1: Breakdown of the ‘Medical corpus’ - texts produced by UK medical organisations** | | | | | | |
| --- | --- | --- | --- | --- | --- | --- |
| **Organisation** | **Text** | **Date accessed** | **Date published/reviewed** | **Text-type** | **URL** | **Primary audience** |
| **NICE (National Institute for Health and Care Excellence)** | Ectopic pregnancy and miscarriage: diagnosis and initial management NG126 | April 2025 | 17 April 2019 last updated: August 2023 | guidelines | [Ectopic pregnancy and miscarriage: diagnosis and initial management \| NICE](https://www.nice.org.uk/guidance/ng126/resources/ectopic-pregnancy-and-miscarriage-diagnosis-and-initial-management-pdf-66141662244037) | Healthcare professionals, commissioners or trainees |
|  | Antenatal care NG201 | April 2025 | 19 August 2021 last updated: December 2024 | guidelines | [Antenatal care \| NICE](https://www.nice.org.uk/guidance/ng201/resources/antenatal-care-pdf-66143709695941) | Healthcare professionals, commissioners or trainees |
|  | Twin and triplet pregnancy NG137 | April 2025 | 04 September 2019 last updated: 09 April 2024 | guidelines | [Twin and triplet pregnancy \| NICE](https://www.nice.org.uk/guidance/ng137/resources/twin-and-triplet-pregnancy-pdf-66141724389829) | Healthcare professionals, commissioners or trainees |
|  | Antenatal and postnatal mental health: clinical management and service guidance CG192 | April 2025 | 17 December 2014 last updated February 2020 | guidelines | [Antenatal and postnatal mental health- clinical management and service guidance – NICE](https://www.nice.org.uk/guidance/cg192/resources/antenatal-and-postnatal-mental-health-clinical-management-and-service-guidance-pdf-35109869806789) | Healthcare professionals, commissioners or trainees |
|  | Ectopic pregnancy and miscarriage QS69 | April 2025 | 10 September 2014 | quality standards | [Ectopic pregnancy and miscarriage \| NICE](https://www.nice.org.uk/guidance/qs69/resources/ectopic-pregnancy-and-miscarriage-pdf-2098796040133) | Healthcare professionals, commissioners or trainees |
| **NHS (National Health Service)** | Overview of Miscarriage on the main NHS page | April 2025 | 9 March 2022 | Patient-facing, informational web pages | [Miscarriage - NHS](https://www.nhs.uk/conditions/miscarriage/) | General public, patients, caregivers |
|  | Overview of Ectopic Pregnancy on the main NHS page | April 2025 | 23 August 2022 | Patient-facing, informational web pages | [Ectopic pregnancy - NHS](https://www.nhs.uk/conditions/ectopic-pregnancy/) | General public, patients, caregivers |
|  | Overview of Molar Pregnancy on the main NHS page | April 2025 | 19 June 2023 | Patient-facing, informational web pages | [Molar pregnancy - NHS](https://www.nhs.uk/conditions/molar-pregnancy/) | General public, patients, caregivers |
|  | Overview of Stillbirth on the main NHS page | April 2025 | 24 April 2024 | Patient-facing, informational web pages | [Stillbirth - NHS](https://www.nhs.uk/conditions/stillbirth/) | General public, patients, caregivers |
|  | When pregnancy goes wrong | April 2025 | 30 April 2024 | Patient-facing, informational web pages | [When pregnancy goes wrong - NHS](https://www.nhs.uk/pregnancy/support/when-pregnancy-goes-wrong/) | General public, patients, caregivers |
|  | Termination for fetal anomaly | April 2025 | 25 July 2024 | Patient-facing, informational web pages | [Termination for fetal anomaly - NHS](https://www.nhs.uk/pregnancy/support/termination-for-fetal-anomaly/) | General public, patients, caregivers |
|  | What you can do to reduce the risk of stillbirth | April 2025 | 19 July 2024 | Patient-facing, informational web pages | [What you can do to reduce the risk of stillbirth - NHS](https://www.nhs.uk/pregnancy/keeping-well/reducing-the-risk-of-stillbirth/) | General public, patients, caregivers |
|  | If antenatal screening tests find something | April 2025 | 29 November 2022 | Patient-facing, informational web pages | [If antenatal screening tests find something - NHS](https://www.nhs.uk/pregnancy/your-pregnancy-care/if-antenatal-screening-tests-find-something/) | General public, patients, caregivers |
| **Royal College of Obstetricians and Gynaecologists** | Termination of pregnancy for fetal abnormality | April 2025 | May 2010 | Clinical guidelines | [Termination of Pregnancy for Fetal Abnormality](https://www.rcog.org.uk/media/21lfvl0e/terminationpregnancyreport18may2010.pdf) | Healthcare professionals, commissioners or trainees |
|  | Recurrent Miscarriage Guideline No. 17 | April 2025 | June 2023 | Clinical guidelines | [Recurrent Miscarriage (Green-top Guideline No. 17) \| RCOG](https://www.rcog.org.uk/guidance/browse-all-guidance/green-top-guidelines/recurrent-miscarriage-green-top-guideline-no-17/) | Healthcare professionals, commissioners or trainees |
|  | Diagnosis and Management of Ectopic Pregnancy Guideline No. 21 | April 2025 | November 2016 | Clinical guidelines | [Diagnosis and Management of Ectopic Pregnancy (Green-top Guideline No. 21) \| RCOG](https://www.rcog.org.uk/guidance/browse-all-guidance/green-top-guidelines/diagnosis-and-management-of-ectopic-pregnancy-green-top-guideline-no-21/) | Healthcare professionals, commissioners or trainees |
|  | Care of late intrauterine fetal death and stillbirth Guideline No. 55 | April 2025 | October 2024 | Clinical guidelines | [Care of late intrauterine fetal death and stillbirth (Green-top Guideline No. 55) \| RCOG](https://www.rcog.org.uk/guidance/browse-all-guidance/green-top-guidelines/care-of-late-intrauterine-fetal-death-and-stillbirth-green-top-guideline-no-55/) | Healthcare professionals, commissioners or trainees |
|  | Surgical Management of Miscarriage and Removal of Persistent Placental or Fetal Remains (Consent Advice No. 10) | April 2025 | January 2018 | Consent Advice | [Surgical Management of Miscarriage and Removal of Persistent Placental or Fetal Remains](https://www.rcog.org.uk/media/mslbkndx/consent-advice-10-surgical-management-of-miscarriage.pdf) | Healthcare professionals, commissioners or trainees |
|  | Early miscarriage | April 2025 | September 2016 | patient information | [Early miscarriage \| RCOG](https://www.rcog.org.uk/for-the-public/browse-our-patient-information/early-miscarriage/) | General public, patients, caregivers |
|  | Recurrent Miscarriage | April 2025 | June 2023 | patient information | [Recurrent miscarriage \| RCOG](https://www.rcog.org.uk/for-the-public/browse-our-patient-information/recurrent-miscarriage/) | General public, patients, caregivers |
|  | Ectopic pregnancy | May 2025 | November 2017 | patient information | [Ectopic pregnancy \| RCOG](https://www.rcog.org.uk/for-the-public/browse-our-patient-information/ectopic-pregnancy/) | General public, patients, caregivers |
|  | Recovering from surgical management of a miscarriage | May 2025 | October 2015 | patient information | [Recovering from surgical management of a miscarriage \| RCOG](https://www.rcog.org.uk/for-the-public/browse-our-patient-information/recovering-from-surgical-management-of-a-miscarriage/) | General public, patients, caregivers |
|  | Molar pregnancy and gestational trophoblastic disease | May 2025 | September 2020 | patient information | [Molar pregnancy and gestational trophoblastic disease \| RCOG](https://www.rcog.org.uk/for-the-public/browse-our-patient-information/molar-pregnancy-and-gestational-trophoblastic-disease/) | General public, patients, caregivers |
|  | When your baby dies before birth | May 2025 | July 2012,  last updated October 2024 | patient information | [When your baby dies before birth \| RCOG](https://www.rcog.org.uk/for-the-public/browse-our-patient-information/when-your-baby-dies-before-birth/) | General public, patients, caregivers |
|  | Registration of Stillbirths and Certification for Pregnancy Loss Before 24 Weeks of Gestation (Good Practice No. 4) | May 2025 | January 2005 | best practice paper | [Registration of Stillbirths and Certification for Pregnancy Loss Before 24 Weeks of Gestation (Good Practice No. 4) \| RCOG](https://www.rcog.org.uk/guidance/browse-all-guidance/good-practice-papers/registration-of-stillbirths-and-certification-for-pregnancy-loss-before-24-weeks-of-gestation-good-practice-no-4/) | Healthcare professionals, commissioners or trainees |
|  | Subspecialty Training Reproductive Medicine Definitive Document | June 2026 | March 2024 | training curriculum | https://www.gmc-uk.org/cdn/documents/rm-sst-2024---definitive-document---final-march-2024_pdf-106366463.pdf | Healthcare professionals, commissioners or trainees |
|  | Special Interest Training obstretrics and Gynaecology | June 2026 | March 2024 | training curriculum | https://www.gmc-uk.org/cdn/documents/special-interest-training-2024---definitive-document---final-march-2024_pdf-106362539.pdf | Healthcare professionals, commissioners or trainees |
| **Royal College of General Practitioners** | Maternity and reproductive health | June 2026 | August 2025 | Curriculum topic guides | https://www.gmc-uk.org/cdn/documents/topic-guides-2025_pdf-110256640.pdf | Healthcare professionals, commissioners or trainees |
| **Royal College of Pathologists** | Curriculum for speciality training in paediatric and perinatal | June 2026 | August 2021 | training curriculum | https://www.gmc-uk.org/cdn/documents/2021-paediatric-and-perinatal-pathology-curriculum_pdf-87181858.pdf | Healthcare professionals, commissioners or trainees |
| **NUPAS (National Unplanned Pregnancy Advisory Service)** | Termination for medical reasons | May 2025 | March 2023 | patient information | [Terminating a Pregnancy For Medical Reasons \| NUPAS](https://www.nupas.co.uk/abortion-care/termination-for-medical-reasons/) | General public, patients, caregivers |
| **MBRRACE-UK (Mothers and Babies: Reducing Risk through Audits and Confidential Enquiries across the UK)** | Perinatal Mortality Surveillance: UK perinatal deaths of babies born in 2023 | May 2025 | May 2025 | statistical and audit report | [MBRRACE-UK Perinatal Mortality Surveillance](https://timms.le.ac.uk/mbrrace-uk-perinatal-mortality/surveillance/) | Healthcare professionals, commissioners or trainees |

**Table 2: Breakdown of the ‘Charity corpus’ - texts produced by UK based charities**

| **Organisation** | **Text** | **Date accessed** | **Date published/reviewed** | **Text- type** | **URL** | **Primary audience** |
| --- | --- | --- | --- | --- | --- | --- |
| **Tommy's** | Miscarriage information and support | May 2025 | reviewed in 2022-2024 | patient information | [Miscarriage information and support \| Tommy's](https://www.tommys.org/baby-loss-support/miscarriage-information-and-support) | General public, patients, caregivers |
|  | Ectopic Pregnancy | May 2025 | reviewed in February 2024 | patient information | [Ectopic Pregnancy: Signs, Treatment and Support](https://www.tommys.org/baby-loss-support/ectopic-pregnancy-information-support) | General public, patients, caregivers |
|  | Molar Pregnancy | May 2025 | reviewed in February 2025 | patient information | [Molar Pregnancy: Definition, Types, Symptoms & Risk Factors](https://www.tommys.org/baby-loss-support/molar-pregnancy-information-support) | General public, patients, caregivers |
|  | Termination for Medical Reasons | May 2025 | May 2023 | patient information | [Terminating a Pregnancy for Medical Reasons (TFMR) \| Tommy's](https://www.tommys.org/baby-loss-support/tfmr-terminating-pregnancy-medical-reasons) | General public, patients, caregivers |
|  | Stillbirth | May 2025 | reviewed in 2022-2025 | patient information | [Stillbirth information and support \| Tommy's](https://www.tommys.org/baby-loss-support/stillbirth-information-and-support) | General public, patients, caregivers |
| **Petals** | Miscarriage | June 2025 | June 2025 | patient information | [Miscarriage - Petals Charity](https://www.petalscharity.org/about-baby-loss/aboutmiscarriage/) | General public, patients, caregivers |
|  | Recurrent miscarriage | June 2025 | June 2025 | patient information | [Recurrent Miscarriage - Petals Charity](https://www.petalscharity.org/about-baby-loss/aboutrecurrentmiscarriage/) | General public, patients, caregivers |
|  | Termination for Medical Reasons | June 2025 | June 2025 | patient information | [Termination For Medical Reasons (TFMR) - Petals Charity](https://www.petalscharity.org/about-baby-loss/abouttfmr/) | General public, patients, caregivers |
|  | Stillbirth | June 2025 | June 2025 | patient information | [Stillbirth - Petals Charity](https://www.petalscharity.org/about-baby-loss/aboutstillbirth/) | General public, patients, caregivers |
|  | Petals leaflet | June 2025 | June 2025 | patient information | [Providing FREE, specialist counselling to individuals and couples after pregnancy and baby loss.](https://www.petalscharity.org/wp-content/uploads/Petals-tri-fold-leaflet.pdf) | General public, patients, caregivers |
| **Miscarriage Association** | Miscarriage | May 2025 | September 2016 | patient information | [The Miscarriage Association](https://www.miscarriageassociation.org.uk/information/miscarriage/) | General public, patients, caregivers |
|  | Ectopic Pregnancy | May 2025 | September 2016 | patient information | [Ectopic pregnancy - The Miscarriage Association](https://www.miscarriageassociation.org.uk/information/ectopic-pregnancy/) | General public, patients, caregivers |
|  | Molar Pregnancy | May 2025 | September 2016 | patient information | [Molar pregnancy - The Miscarriage Association](https://www.miscarriageassociation.org.uk/information/molar-pregnancy/) | General public, patients, caregivers |
|  | Pregnancy Loss within Black and Black Mixed Heritage communities | May 2025 | May 2025 | patient information | [Pregnancy loss within Black and Black mixed heritage communities - The Miscarriage Association](https://www.miscarriageassociation.org.uk/information/pregnancy-loss-within-black-and-black-mixed-heritage-communities/) | General public, patients, caregivers |
|  | Pregnancy Loss within the LGBTQIA community | May 2025 | June 2024 | patient information | [Pregnancy loss within the LGBTQIA community - The Miscarriage Association](https://www.miscarriageassociation.org.uk/information/pregnancy-loss-lgbtqia-community/) | General public, patients, caregivers |
|  | For Health Professionals | May 2025 | September 2016 | patient information | [For health professionals - The Miscarriage Association](https://www.miscarriageassociation.org.uk/information/for-health-professionals/) | Healthcare professionals, commissioners or trainees |
|  | Worried About Pregnancy Loss? | May 2025 | September 2016 | patient information | [Worried about pregnancy loss? - The Miscarriage Association](https://www.miscarriageassociation.org.uk/information/worried-about-pregnancy-loss/) | General public, patients, caregivers |
|  | A Range of Emotions | May 2025 | January 2017 | patient information -- mental health | [Emotions after a miscarriage](https://www.miscarriageassociation.org.uk/your-feelings/common-feelings/) | General public, patients, caregivers |
|  | Special Circumstances | May 2025 | September 2016, updated January 2023 | patient information -- mental health | [Special circumstances - The Miscarriage Association](https://www.miscarriageassociation.org.uk/your-feelings/special-circumstances/) | General public, patients, caregivers |
|  | Your Mental Health | May 2025 | April 2019, updated September 2022 | patient information -- mental health | [Your mental health - The Miscarriage Association](https://www.miscarriageassociation.org.uk/your-feelings/your-mental-health/) | General public, patients, caregivers |
| **Sands** | Support for all types of baby loss | May 2025 | October 2024 | Patient information | [Support for all types of baby loss \| Sands - Saving babies' lives. Supporting bereaved families.](https://www.sands.org.uk/support-all-types-baby-loss) | General public, patients, caregivers |
|  | Understanding Why Your Baby Died | May 2025 | March 2025 | Patient information | [Understanding why your baby died \| Sands - Saving babies' lives. Supporting bereaved families.](https://www.sands.org.uk/understanding-why-your-baby-died) | General public, patients, caregivers |
|  | Giving Birth To Your Baby | May 2025 | March 2023 | Patient-facing booklet (guide) | [Giving birth to your baby](https://www.sands.org.uk/sites/default/files/Giving_Birth_To_Your_Baby_March_2023.pdf) | General public, patients, caregivers |
|  | Information and Support for Partners | May 2025 | March 2023 | Patient-facing booklet (guide) | [Information and support for partners](https://www.sands.org.uk/sites/default/files/Information_And_Support_For_Partners_March_2023.pdf) | General public, patients, caregivers |
|  | Remembering Your Baby | May 2025 | March 2023 | Patient-facing booklet (guide) | [Remembering your baby](https://www.sands.org.uk/sites/default/files/Remembering_Your_Baby_March_2023.pdf) | General public, patients, caregivers |
|  | Understanding why your baby died | May 2025 | March 2023 | Patient-facing booklet (guide) | [Understanding why your baby died](https://www.sands.org.uk/sites/default/files/Understanding_why_your_baby_died.pdf) | General public, patients, caregivers |
|  | Ending a pregnancy for medical reasons (TFMR) | May 2025 | March 2023 | Patient-facing booklet (guide) | [Ending a pregnancy for medical reasons (TFMR)](https://www.sands.org.uk/sites/default/files/Ending_A_Pregnancy_For_Medical_Reasons_(TFMR)_March_2023.pdf) | General public, patients, caregivers |
|  | Saying goodbye to your baby | May 2025 | March 2023 | Patient-facing booklet (guide) | [Saying goodbye to your baby](https://www.sands.org.uk/sites/default/files/SandsBereavementBook_SayingGoodbye_Nov24_Digital.pdf) | General public, patients, caregivers |
|  | Physical and emotional challenges | May 2025 | March 2023 | Patient-facing booklet (guide) | [Physical and emotional challenges](https://www.sands.org.uk/sites/default/files/Physical_And_Emotional_Challenges_March_2023.pdf) | General public, patients, caregivers |
| **BPAS (British Pregnancy Advisory Service)** | Miscarriage care | May 2025 | September 2023 | Patient information | [Miscarriage care \| BPAS](https://www.bpas.org/more-services-information/pregnancy-miscarriage/miscarriage-care/) | General public, patients, caregivers |
|  | Termination for fetal anomaly | May 2025 | September 2023 | Patient information | [Termination for fetal anomaly - Pregnancy & miscarriage](https://www.bpas.org/more-services-information/pregnancy-miscarriage/termination-for-fetal-anomaly/) | General public, patients, caregivers |
| **The Ectopic Pregnancy Trust** | What is an ectopic pregnancy? | May 2025 | November 2023 | Patient information | [What Is An Ectopic Pregnancy?](https://ectopic.org.uk/what-is-an-ectopic-pregnancy) | General public, patients, caregivers |
|  | Symptoms of an ectopic pregnancy | May 2025 | June 2021 | Patient information | [Ectopic Pregnancy Signs And Symptoms](https://ectopic.org.uk/ectopic-pregnancy-symptoms) | General public, patients, caregivers |
|  | Ectopic Pregnancy Diagnosis | May 2025 | June 2021 | Patient information | [Ectopic Pregnancy Diagnosis](https://ectopic.org.uk/diagnosing-an-ectopic-pregnancy) | General public, patients, caregivers |
|  | Treating an Ectopic Pregnancy | May 2025 | June 2021 -- April 2023 | Patient information | [Treating An Ectopic Pregnancy](https://ectopic.org.uk/treating-an-ectopic-pregnancy) | General public, patients, caregivers |
|  | Reasons for an Ectopic Pregnancy | May 2025 | April 2023 | Patient information | [Reasons Why An Ectopic Pregnancy Can Occur](https://ectopic.org.uk/reasons-for-an-ectopic-pregnancy) | General public, patients, caregivers |
|  | Physical Recovery | May 2025 | June 2021 | Patient information | [Physical Recovery From An Ectopic Pregnancy](https://ectopic.org.uk/physical-recovery) | General public, patients, caregivers |
|  | Emotional Recovery | May 2025 | June 2021 | Patient information | [Emotional Recovery From An Ectopic Pregnancy](https://ectopic.org.uk/emotional-recovery) | General public, patients, caregivers |
| **ARC (Antenatal Results and Choices)** | Introduction | May 2025 | Information not available | Patient information | [Tests explained](https://www.arc-uk.org/tests-explained/) | General public, patients, caregivers |
|  | Screening Tests | May 2025 | Information not available | Patient information | [Screening for Down’s syndrome, Edwards’ syndrome and Patau’s syndrome – Antenatal Results and Choices (ARC)](https://www.arc-uk.org/tests-explained/screening-for-downs-syndrome-edwards-syndrome-and-pataus-syndrome/) | General public, patients, caregivers |
|  | Diagnostic Tests | May 2025 | Information not available | Patient information | [Diagnostic tests](https://www.arc-uk.org/tests-explained/diagnostic-tests/) | General public, patients, caregivers |
|  | Genetic Tests | May 2025 | Information not available | Patient information | [Prenatal genetic screening tests](https://www.arc-uk.org/tests-explained/prenatal-genetic-screening-tests/) | General public, patients, caregivers |
|  | After Diagnosis | May 2025 | Information not available | Patient information | [For parents](https://www.arc-uk.org/for-parents/) | General public, patients, caregivers |
|  | Dealing With Loss | May 2025 | Information not available | Patient information | [Our bereavement support – Antenatal Results and Choices (ARC)](https://www.arc-uk.org/for-parents/arc-bereavement-support/) | General public, patients, caregivers |
|  | Parent-centred Care Guidelines | June 2025 | Information not available | Guidelines | [Parent-centred Care Guidelines](https://www.arc-uk.org/for-professionals/parent-centred-care-guidelines/) | Healthcare professionals, commissioners or trainees |
|  | Supporting you through your pregnancy | June 2025 | reviewed June 2023 | Patient information | https://www.arc-uk.org/publication/supporting-you-through-your-pregnancy/ | General public, patients, caregivers |
|  | Another pregnancy | June 2025 | Information not available | Patient information | https://www.arc-uk.org/for-parents/another-pregnancy/ | General public, patients, caregivers |
| **Molar Pregnancy support and information** | Molar Pregnancy Information | June 2025 | April 2016 | Patient information | [Molar Pregnancy Sheffield Information & Support](https://molarpregnancy.co.uk/information/) | General public, patients, caregivers |
