## Supplemental material: Appendix 2 for "Communication without dialogue? Framing patient–professional interaction in UK institutional discourse on pregnancy loss"

**Table 1: Positive keywords in Medical corpus (120,383 words) compared to Charity corpus (224,481 words)**

Excluded zeros in column 1, min. freq 1, LL greater than 6.63, sorted by effect size (LogRatio), sample of top 150 results:

|  | **Item O1 %1 O2 %2 LL LogRatio** |
| --- | --- |
| 1. | recommendation 340 0.24 2 0.00 + 701.60 8.34 |
| 2. | rationale 84 0.06 1 0.00 + 168.96 7.32 |
| 3. | committee 168 0.12 2 0.00 + 337.91 7.32 |
| 4. | postmortem 67 0.05 1 0.00 + 133.18 6.99 |
| 5. | serum 105 0.07 2 0.00 + 205.54 6.64 |
| 6. | 2012 92 0.06 2 0.00 + 178.36 6.45 |
| 7. | triplet 91 0.06 2 0.00 + 176.28 6.44 |
| 8. | Recommendations 215 0.15 5 0.00 + 414.61 6.35 |
| 9. | systematic 42 0.03 1 0.00 + 80.84 6.32 |
| 10. | monochorionic 41 0.03 1 0.00 + 78.76 6.29 |
| 11. | 1,000 36 0.03 1 0.00 + 68.36 6.10 |
| 12. | table 35 0.02 1 0.00 + 66.28 6.06 |
| 13. | systemic 32 0.02 1 0.00 + 60.07 5.93 |
| 14. | trial 31 0.02 1 0.00 + 58.00 5.88 |
| 15. | rates 186 0.13 6 0.00 + 348.00 5.88 |
| 16. | 95% 87 0.06 3 0.00 + 161.61 5.79 |
| 17. | morbidity 28 0.02 1 0.00 + 51.81 5.73 |
| 18. | fetuses 25 0.02 1 0.00 + 45.64 5.57 |
| 19. | &lsqb; 332 0.23 14 0.01 + 602.02 5.50 |
| 20. | &rsqb; 331 0.23 14 0.01 + 599.97 5.49 |
| 21. | population 23 0.02 1 0.00 + 41.54 5.45 |
| 22. | mortality 125 0.09 6 0.00 + 222.70 5.31 |
| 23. | births 104 0.07 5 0.00 + 185.24 5.31 |
| 24. | resection 20 0.01 1 0.00 + 35.42 5.25 |
| 25. | al 20 0.01 1 0.00 + 35.42 5.25 |
| 26. | guideline 99 0.07 5 0.00 + 175.07 5.23 |
| 27. | states 19 0.01 1 0.00 + 33.39 5.18 |
| 28. | secondary 19 0.01 1 0.00 + 33.39 5.18 |
| 29. | adnexal 19 0.01 1 0.00 + 33.39 5.18 |
| 30. | proportion 37 0.03 2 0.00 + 64.75 5.14 |
| 31. | 2019 73 0.05 4 0.00 + 127.48 5.12 |
| 32. | gtd 18 0.01 1 0.00 + 31.36 5.10 |
| 33. | classification 18 0.01 1 0.00 + 31.36 5.10 |
| 34. | importance 17 0.01 1 0.00 + 29.34 5.01 |
| 35. | severity 16 0.01 1 0.00 + 27.33 4.93 |
| 36. | interval 16 0.01 1 0.00 + 27.33 4.93 |
| 37. | dichorionic 16 0.01 1 0.00 + 27.33 4.93 |
| 38. | decreased 16 0.01 1 0.00 + 27.33 4.93 |
| 39. | aneuploidy 16 0.01 1 0.00 + 27.33 4.93 |
| 40. | strength 79 0.05 5 0.00 + 134.65 4.91 |
| 41. | data 106 0.07 7 0.00 + 179.28 4.85 |
| 42. | terminated 15 0.01 1 0.00 + 25.33 4.83 |
| 43. | amended 45 0.03 3 0.00 + 75.98 4.83 |
| 44. | 2014 15 0.01 1 0.00 + 25.33 4.83 |
| 45. | 2- 15 0.01 1 0.00 + 25.33 4.83 |
| 46. | 2023 59 0.04 4 0.00 + 99.30 4.81 |
| 47. | mutation 14 0.01 1 0.00 + 23.33 4.73 |
| 48. | ethnic 14 0.01 1 0.00 + 23.33 4.73 |
| 49. | evaluation 13 0.01 1 0.00 + 21.34 4.63 |
| 50. | tract 12 0.01 1 0.00 + 19.36 4.51 |
| 51. | no. 12 0.01 1 0.00 + 19.36 4.51 |
| 52. | controls 12 0.01 1 0.00 + 19.36 4.51 |
| 53. | analgesia 12 0.01 1 0.00 + 19.36 4.51 |
| 54. | overall 35 0.02 3 0.00 + 56.12 4.47 |
| 55. | agreed 68 0.05 6 0.00 + 108.32 4.43 |
| 56. | studies 144 0.10 13 0.00 + 228.15 4.40 |
| 57. | status 11 0.01 1 0.00 + 17.40 4.39 |
| 58. | determined 11 0.01 1 0.00 + 17.40 4.39 |
| 59. | collected 22 0.02 2 0.00 + 34.80 4.39 |
| 60. | biochemical 22 0.02 2 0.00 + 34.80 4.39 |
| 61. | assessing 11 0.01 1 0.00 + 17.40 4.39 |
| 62. | 1(1) 11 0.01 1 0.00 + 17.40 4.39 |
| 63. | outcomes 108 0.08 10 0.00 + 170.08 4.36 |
| 64. | laparoscopic 32 0.02 3 0.00 + 50.24 4.34 |
| 65. | undertaken 21 0.01 2 0.00 + 32.85 4.32 |
| 66. | detection 21 0.01 2 0.00 + 32.85 4.32 |
| 67. | marked 10 0.01 1 0.00 + 15.45 4.25 |
| 68. | indications 10 0.01 1 0.00 + 15.45 4.25 |
| 69. | figure 20 0.01 2 0.00 + 30.90 4.25 |
| 70. | consensus 10 0.01 1 0.00 + 15.45 4.25 |
| 71. | anaesthesia 10 0.01 1 0.00 + 15.45 4.25 |
| 72. | abortions 10 0.01 1 0.00 + 15.45 4.25 |
| 73. | substantial 19 0.01 2 0.00 + 28.97 4.18 |
| 74. | practitioners 19 0.01 2 0.00 + 28.97 4.18 |
| 75. | failure 19 0.01 2 0.00 + 28.97 4.18 |
| 76. | evidence 497 0.35 52 0.02 + 758.91 4.18 |
| 77. | concluded 19 0.01 2 0.00 + 28.97 4.18 |
| 78. | context 28 0.02 3 0.00 + 42.48 4.15 |
| 79. | adverse 55 0.04 6 0.00 + 83.04 4.12 |
| 80. | practice 163 0.11 18 0.01 + 245.29 4.11 |
| 81. | resulted 9 0.01 1 0.00 + 13.52 4.10 |
| 82. | q 9 0.01 1 0.00 + 13.52 4.10 |
| 83. | prevalence 9 0.01 1 0.00 + 13.52 4.10 |
| 84. | optimal 18 0.01 2 0.00 + 27.04 4.10 |
| 85. | interpretation 9 0.01 1 0.00 + 13.52 4.10 |
| 86. | conducted 9 0.01 1 0.00 + 13.52 4.10 |
| 87. | categories 9 0.01 1 0.00 + 13.52 4.10 |
| 88. | anticoagulant 9 0.01 1 0.00 + 13.52 4.10 |
| 89. | 3d 9 0.01 1 0.00 + 13.52 4.10 |
| 90. | intrapartum 26 0.02 3 0.00 + 38.64 4.04 |
| 91. | 1000 26 0.02 3 0.00 + 38.64 4.04 |
| 92. | endometrial 17 0.01 2 0.00 + 25.13 4.01 |
| 93. | improved 25 0.02 3 0.00 + 36.74 3.99 |
| 94. | account 33 0.02 4 0.00 + 48.35 3.97 |
| 95. | unbalanced 8 0.01 1 0.00 + 11.61 3.93 |
| 96. | suspicion 16 0.01 2 0.00 + 23.22 3.93 |
| 97. | proposed 8 0.01 1 0.00 + 11.61 3.93 |
| 98. | obesity 8 0.01 1 0.00 + 11.61 3.93 |
| 99. | leiden 8 0.01 1 0.00 + 11.61 3.93 |
| 100. | frequent 8 0.01 1 0.00 + 11.61 3.93 |
| 101. | diameter 8 0.01 1 0.00 + 11.61 3.93 |
| 102. | clinically 24 0.02 3 0.00 + 34.84 3.93 |
| 103. | chronic 8 0.01 1 0.00 + 11.61 3.93 |
| 104. | assessments 8 0.01 1 0.00 + 11.61 3.93 |
| 105. | ages 8 0.01 1 0.00 + 11.61 3.93 |
| 106. | 34 8 0.01 1 0.00 + 11.61 3.93 |
| 107. | 24th 8 0.01 1 0.00 + 11.61 3.93 |
| 108. | perinatal 110 0.08 14 0.01 + 158.78 3.90 |
| 109. | expertise 31 0.02 4 0.00 + 44.56 3.88 |
| 110. | measures 23 0.02 3 0.00 + 32.95 3.87 |
| 111. | compared 77 0.05 10 0.00 + 110.45 3.87 |
| 112. | subsequent 76 0.05 10 0.00 + 108.56 3.85 |
| 113. | trophoblast 15 0.01 2 0.00 + 21.34 3.83 |
| 114. | survival 15 0.01 2 0.00 + 21.34 3.83 |
| 115. | anomalies 82 0.06 11 0.00 + 116.41 3.83 |
| 116. | accuracy 15 0.01 2 0.00 + 21.34 3.83 |
| 117. | preterm 81 0.06 11 0.00 + 114.53 3.81 |
| 118. | 2021 22 0.02 3 0.00 + 31.07 3.80 |
| 119. | statement 29 0.02 4 0.00 + 40.80 3.79 |
| 120. | intrauterine 80 0.06 11 0.00 + 112.66 3.79 |
| 121. | abnormality 87 0.06 12 0.00 + 122.39 3.79 |
| 122. | undiagnosed 7 0.00 1 0.00 + 9.73 3.73 |
| 123. | tsh 14 0.01 2 0.00 + 19.46 3.73 |
| 124. | target 7 0.00 1 0.00 + 9.73 3.73 |
| 125. | septate 14 0.01 2 0.00 + 19.46 3.73 |
| 126. | searched 7 0.00 1 0.00 + 9.73 3.73 |
| 127. | reduction 35 0.02 5 0.00 + 48.66 3.73 |
| 128. | raw 7 0.00 1 0.00 + 9.73 3.73 |
| 129. | prompt 14 0.01 2 0.00 + 19.46 3.73 |
| 130. | presenting 7 0.00 1 0.00 + 9.73 3.73 |
| 131. | mandatory 7 0.00 1 0.00 + 9.73 3.73 |
| 132. | intramuscular 7 0.00 1 0.00 + 9.73 3.73 |
| 133. | hydrops 7 0.00 1 0.00 + 9.73 3.73 |
| 134. | handicapped 7 0.00 1 0.00 + 9.73 3.73 |
| 135. | dysfunction 7 0.00 1 0.00 + 9.73 3.73 |
| 136. | childhood 7 0.00 1 0.00 + 9.73 3.73 |
| 137. | basis 14 0.01 2 0.00 + 19.46 3.73 |
| 138. | audit 14 0.01 2 0.00 + 19.46 3.73 |
| 139. | administration 14 0.01 2 0.00 + 19.46 3.73 |
| 140. | incidence 41 0.03 6 0.00 + 56.53 3.70 |
| 141. | total 27 0.02 4 0.00 + 37.07 3.68 |
| 142. | thrombophilia 26 0.02 4 0.00 + 35.22 3.63 |
| 143. | summary 13 0.01 2 0.00 + 17.61 3.63 |
| 144. | reviewed 13 0.01 2 0.00 + 17.61 3.63 |
| 145. | obtained 13 0.01 2 0.00 + 17.61 3.63 |
| 146. | minimum 13 0.01 2 0.00 + 17.61 3.63 |
| 147. | features 26 0.02 4 0.00 + 35.22 3.63 |
| 148. | 2022 26 0.02 4 0.00 + 35.22 3.63 |
| 149. | woman 198 0.14 31 0.01 + 266.51 3.60 |
| 150. | reported 89 0.06 14 0.01 + 119.59 3.60 |

**Table 2: Positive** **keywords in Charity corpus (224,481 words) compared to Medical corpus (120,383 words)**

Excluded zeros in column 1, min. freq 1, LL greater than 6.63, sorted by effect size (LogRatio), sample of top 150 results:

**Item	O1	%1	O2	%2	LL LogRatio**

| 1. | us 216 0.08 1 0.00 + 171.90 6.83 |
| --- | --- |
| 2. | say 140 0.05 1 0.00 + 108.54 6.20 |
| 3. | talking 132 0.05 2 0.00 + 95.01 5.12 |
| 4. | nipt 65 0.02 1 0.00 + 46.69 5.09 |
| 5. | kind 63 0.02 1 0.00 + 45.06 5.05 |
| 6. | our 478 0.17 9 0.01 + 333.42 4.80 |
| 7. | thing 50 0.02 1 0.00 + 34.54 4.72 |
| 8. | his 50 0.02 1 0.00 + 34.54 4.72 |
| 9. | upset 49 0.02 1 0.00 + 33.73 4.69 |
| 10. | perhaps 89 0.03 2 0.00 + 60.24 4.55 |
| 11. | birthing 89 0.03 2 0.00 + 60.24 4.55 |
| 12. | here 266 0.10 6 0.00 + 179.92 4.54 |
| 13. | him 43 0.02 1 0.00 + 28.92 4.50 |
| 14. | &quot; 505 0.18 12 0.01 + 338.26 4.47 |
| 15. | everyone 77 0.03 2 0.00 + 50.67 4.34 |
| 16. | hard 141 0.05 4 0.00 + 91.06 4.21 |
| 17. | pages 34 0.01 1 0.00 + 21.78 4.16 |
| 18. | ashes 34 0.01 1 0.00 + 21.78 4.16 |
| 19. | thoughts 65 0.02 2 0.00 + 41.20 4.09 |
| 20. | quite 32 0.01 1 0.00 + 20.21 4.07 |
| 21. | comes 32 0.01 1 0.00 + 20.21 4.07 |
| 22. | ceremony 32 0.01 1 0.00 + 20.21 4.07 |
| 23. | me 159 0.06 5 0.00 + 100.26 4.06 |
| 24. | helped 63 0.02 2 0.00 + 39.63 4.05 |
| 25. | cope 93 0.03 3 0.00 + 58.28 4.03 |
| 26. | page 120 0.04 4 0.00 + 74.58 3.98 |
| 27. | knowing 30 0.01 1 0.00 + 18.65 3.98 |
| 28. | gone 30 0.01 1 0.00 + 18.65 3.98 |
| 29. | gentle 30 0.01 1 0.00 + 18.65 3.98 |
| 30. | feedback 30 0.01 1 0.00 + 18.65 3.98 |
| 31. | thinking 58 0.02 2 0.00 + 35.73 3.93 |
| 32. | saying 29 0.01 1 0.00 + 17.87 3.93 |
| 33. | journey 28 0.01 1 0.00 + 17.09 3.88 |
| 34. | crematorium 28 0.01 1 0.00 + 17.09 3.88 |
| 35. | 0800 28 0.01 1 0.00 + 17.09 3.88 |
| 36. | 0147 28 0.01 1 0.00 + 17.09 3.88 |
| 37. | tfmr 55 0.02 2 0.00 + 33.41 3.85 |
| 38. | sense 54 0.02 2 0.00 + 32.64 3.83 |
| 39. | pads 27 0.01 1 0.00 + 16.32 3.83 |
| 40. | chemical 27 0.01 1 0.00 + 16.32 3.83 |
| 41. | remember 130 0.05 5 0.00 + 77.73 3.77 |
| 42. | ibuprofen 26 0.01 1 0.00 + 15.55 3.77 |
| 43. | goes 26 0.01 1 0.00 + 15.55 3.77 |
| 44. | frightening 26 0.01 1 0.00 + 15.55 3.77 |
| 45. | happened 152 0.06 6 0.00 + 90.21 3.74 |
| 46. | hear 50 0.02 2 0.00 + 29.56 3.72 |
| 47. | got 25 0.01 1 0.00 + 14.78 3.72 |
| 48. | feels 50 0.02 2 0.00 + 29.56 3.72 |
| 49. | rights 24 0.01 1 0.00 + 14.01 3.66 |
| 50. | idea 47 0.02 2 0.00 + 27.26 3.63 |
| 51. | yourself 162 0.06 7 0.00 + 93.52 3.61 |
| 52. | sadness 46 0.02 2 0.00 + 26.50 3.60 |
| 53. | run 23 0.01 1 0.00 + 13.25 3.60 |
| 54. | bath 23 0.01 1 0.00 + 13.25 3.60 |
| 55. | angry 23 0.01 1 0.00 + 13.25 3.60 |
| 56. | never 67 0.02 3 0.00 + 38.24 3.55 |
| 57. | heal 22 0.01 1 0.00 + 12.49 3.53 |
| 58. | leave 109 0.04 5 0.00 + 61.71 3.52 |
| 59. | vitamin 43 0.02 2 0.00 + 24.23 3.50 |
| 60. | book 64 0.02 3 0.00 + 35.97 3.49 |
| 61. | wear 21 0.01 1 0.00 + 11.74 3.46 |
| 62. | ever 21 0.01 1 0.00 + 11.74 3.46 |
| 63. | emotions 105 0.04 5 0.00 + 58.70 3.46 |
| 64. | break 21 0.01 1 0.00 + 11.74 3.46 |
| 65. | sadly 41 0.01 2 0.00 + 22.73 3.43 |
| 66. | send 20 0.01 1 0.00 + 10.99 3.39 |
| 67. | real 20 0.01 1 0.00 + 10.99 3.39 |
| 68. | mood 20 0.01 1 0.00 + 10.99 3.39 |
| 69. | cvs 40 0.01 2 0.00 + 21.98 3.39 |
| 70. | things 276 0.10 14 0.01 + 150.87 3.37 |
| 71. | tell 155 0.06 8 0.01 + 84.19 3.35 |
| 72. | upsetting 57 0.02 3 0.00 + 30.74 3.32 |
| 73. | telling 19 0.01 1 0.00 + 10.25 3.32 |
| 74. | listen 38 0.01 2 0.00 + 20.49 3.32 |
| 75. | letter 38 0.01 2 0.00 + 20.49 3.32 |
| 76. | knew 19 0.01 1 0.00 + 10.25 3.32 |
| 77. | instructions 19 0.01 1 0.00 + 10.25 3.32 |
| 78. | grave 19 0.01 1 0.00 + 10.25 3.32 |
| 79. | exactly 19 0.01 1 0.00 + 10.25 3.32 |
| 80. | actually 19 0.01 1 0.00 + 10.25 3.32 |
| 81. | comfortable 37 0.01 2 0.00 + 19.75 3.28 |
| 82. | worries 18 0.01 1 0.00 + 9.51 3.24 |
| 83. | understandable 18 0.01 1 0.00 + 9.51 3.24 |
| 84. | sessions 18 0.01 1 0.00 + 9.51 3.24 |
| 85. | sanitary 36 0.01 2 0.00 + 19.01 3.24 |
| 86. | ok 36 0.01 2 0.00 + 19.01 3.24 |
| 87. | month 18 0.01 1 0.00 + 9.51 3.24 |
| 88. | items 18 0.01 1 0.00 + 9.51 3.24 |
| 89. | frightened 18 0.01 1 0.00 + 9.51 3.24 |
| 90. | feelings 252 0.09 14 0.01 + 133.08 3.24 |
| 91. | worried 71 0.03 4 0.00 + 37.29 3.22 |
| 92. | told 123 0.04 7 0.00 + 64.33 3.21 |
| 93. | space 52 0.02 3 0.00 + 27.05 3.19 |
| 94. | memories 52 0.02 3 0.00 + 27.05 3.19 |
| 95. | funeral 173 0.06 10 0.01 + 89.91 3.19 |
| 96. | know 379 0.14 22 0.02 + 196.63 3.18 |
| 97. | please 137 0.05 8 0.01 + 70.90 3.17 |
| 98. | specialises 17 0.01 1 0.00 + 8.77 3.16 |
| 99. | picture 17 0.01 1 0.00 + 8.77 3.16 |
| 100. | huge 17 0.01 1 0.00 + 8.77 3.16 |
| 101. | harder 34 0.01 2 0.00 + 17.54 3.16 |
| 102. | fiscal 17 0.01 1 0.00 + 8.77 3.16 |
| 103. | encourage 17 0.01 1 0.00 + 8.77 3.16 |
| 104. | bit 17 0.01 1 0.00 + 8.77 3.16 |
| 105. | begins 17 0.01 1 0.00 + 8.77 3.16 |
| 106. | hour 33 0.01 2 0.00 + 16.81 3.12 |
| 107. | counsellor 33 0.01 2 0.00 + 16.81 3.12 |
| 108. | tiny 16 0.01 1 0.00 + 8.04 3.07 |
| 109. | note 32 0.01 2 0.00 + 16.09 3.07 |
| 110. | hospitals 96 0.04 6 0.00 + 48.27 3.07 |
| 111. | genome 16 0.01 1 0.00 + 8.04 3.07 |
| 112. | created 16 0.01 1 0.00 + 8.04 3.07 |
| 113. | chosen 16 0.01 1 0.00 + 8.04 3.07 |
| 114. | bottom 16 0.01 1 0.00 + 8.04 3.07 |
| 115. | lost 142 0.05 9 0.01 + 70.95 3.05 |
| 116. | though 47 0.02 3 0.00 + 23.41 3.04 |
| 117. | memory 78 0.03 5 0.00 + 38.78 3.04 |
| 118. | let 78 0.03 5 0.00 + 38.78 3.04 |
| 119. | everything 31 0.01 2 0.00 + 15.37 3.03 |
| 120. | like 320 0.12 21 0.01 + 157.40 3.00 |
| 121. | sonographer 30 0.01 2 0.00 + 14.65 2.98 |
| 122. | sickle 30 0.01 2 0.00 + 14.65 2.98 |
| 123. | relate 15 0.01 1 0.00 + 7.32 2.98 |
| 124. | happening 60 0.02 4 0.00 + 29.30 2.98 |
| 125. | ahead 15 0.01 1 0.00 + 7.32 2.98 |
| 126. | feel 776 0.28 53 0.04 + 374.62 2.94 |
| 127. | wound 29 0.01 2 0.00 + 13.94 2.93 |
| 128. | touch 29 0.01 2 0.00 + 13.94 2.93 |
| 129. | petals 29 0.01 2 0.00 + 13.94 2.93 |
| 130. | n't 623 0.23 43 0.03 + 299.25 2.93 |
| 131. | director 29 0.01 2 0.00 + 13.94 2.93 |
| 132. | right 201 0.07 14 0.01 + 96.12 2.92 |
| 133. | felt 72 0.03 5 0.00 + 34.48 2.92 |
| 134. | hormones 43 0.02 3 0.00 + 20.55 2.91 |
| 135. | worry 70 0.03 5 0.00 + 33.06 2.88 |
| 136. | lining 42 0.02 3 0.00 + 19.84 2.88 |
| 137. | anything 97 0.04 7 0.00 + 45.58 2.87 |
| 138. | box 40 0.01 3 0.00 + 18.43 2.81 |
| 139. | move 52 0.02 4 0.00 + 23.64 2.77 |
| 140. | far 26 0.01 2 0.00 + 11.82 2.77 |
| 141. | through 444 0.16 35 0.02 + 199.23 2.74 |
| 142. | off 63 0.02 5 0.00 + 28.17 2.73 |
| 143. | i 805 0.29 64 0.04 + 359.55 2.73 |
| 144. | again 316 0.12 25 0.02 + 141.52 2.73 |
| 145. | clothes 25 0.01 2 0.00 + 11.13 2.72 |
| 146. | wrong 99 0.04 8 0.01 + 43.82 2.70 |
| 147. | my 296 0.11 24 0.02 + 130.77 2.70 |
| 148. | easy 37 0.01 3 0.00 + 16.35 2.70 |
| 149. | grieve 49 0.02 4 0.00 + 21.57 2.69 |
| 150. | look 181 0.07 15 0.01 + 78.98 2.67 |

**Table 3: Positive keywords in Charity-HP sub-corpus (12,162 words) compared to Medical-HP sub-corpus (91,437 words)**

Excluded zeros in column 1, min. freq 1, LL greater than 6.63, sorted by effect size (LogRatio), sample of top 150 results:

**Item	O1	%1	O2	%2	LL	LogRatio**

| 1. | feedback 11 0.08 1 0.00 + 40.57 6.38 |
| --- | --- |
| 2. | agree 21 0.15 2 0.00 + 77.02 6.31 |
| 3. | here 10 0.07 1 0.00 + 36.46 6.24 |
| 4. | sonographers 8 0.06 1 0.00 + 28.30 5.92 |
| 5. | sessions 7 0.05 1 0.00 + 24.26 5.72 |
| 6. | sector 7 0.05 1 0.00 + 24.26 5.72 |
| 7. | perhaps 7 0.05 1 0.00 + 24.26 5.72 |
| 8. | strongly 13 0.09 2 0.00 + 44.50 5.62 |
| 9. | involvement 6 0.04 1 0.00 + 20.25 5.50 |
| 10. | arc 6 0.04 1 0.00 + 20.25 5.50 |
| 11. | like 11 0.08 2 0.00 + 36.54 5.38 |
| 12. | read 5 0.03 1 0.00 + 16.30 5.24 |
| 13. | questionnaire 5 0.03 1 0.00 + 16.30 5.24 |
| 14. | letter 10 0.07 2 0.00 + 32.60 5.24 |
| 15. | hospitals 5 0.03 1 0.00 + 16.30 5.24 |
| 16. | point 28 0.19 6 0.01 + 89.96 5.14 |
| 17. | know 23 0.16 5 0.00 + 73.67 5.12 |
| 18. | right 4 0.03 1 0.00 + 12.41 4.92 |
| 19. | prepare 4 0.03 1 0.00 + 12.41 4.92 |
| 20. | lip 4 0.03 1 0.00 + 12.41 4.92 |
| 21. | kind 4 0.03 1 0.00 + 12.41 4.92 |
| 22. | call 4 0.03 1 0.00 + 12.41 4.92 |
| 23. | assumptions 4 0.03 1 0.00 + 12.41 4.92 |
| 24. | genomic 7 0.05 2 0.00 + 21.00 4.72 |
| 25. | today 9 0.06 3 0.00 + 25.87 4.50 |
| 26. | think 3 0.02 1 0.00 + 8.62 4.50 |
| 27. | sequencing 6 0.04 2 0.00 + 17.25 4.50 |
| 28. | rights 3 0.02 1 0.00 + 8.62 4.50 |
| 29. | publications 3 0.02 1 0.00 + 8.62 4.50 |
| 30. | pre-test 3 0.02 1 0.00 + 8.62 4.50 |
| 31. | please 9 0.06 3 0.00 + 25.87 4.50 |
| 32. | helpline 3 0.02 1 0.00 + 8.62 4.50 |
| 33. | getting 3 0.02 1 0.00 + 8.62 4.50 |
| 34. | fasp 15 0.10 5 0.00 + 43.12 4.50 |
| 35. | dysplasia 3 0.02 1 0.00 + 8.62 4.50 |
| 36. | contract 3 0.02 1 0.00 + 8.62 4.50 |
| 37. | charity 3 0.02 1 0.00 + 8.62 4.50 |
| 38. | agenesis 3 0.02 1 0.00 + 8.62 4.50 |
| 39. | 23+6 3 0.02 1 0.00 + 8.62 4.50 |
| 40. | your 24 0.17 9 0.01 + 66.56 4.33 |
| 41. | neither 8 0.06 3 0.00 + 22.19 4.33 |
| 42. | sonographer 5 0.03 2 0.00 + 13.58 4.24 |
| 43. | proceed 5 0.03 2 0.00 + 13.58 4.24 |
| 44. | note 5 0.03 2 0.00 + 13.58 4.24 |
| 45. | news 15 0.10 6 0.01 + 40.73 4.24 |
| 46. | book 5 0.03 2 0.00 + 13.58 4.24 |
| 47. | feel 11 0.08 5 0.00 + 28.57 4.05 |
| 48. | want 20 0.14 10 0.01 + 50.12 3.92 |
| 49. | uncertainties 4 0.03 2 0.00 + 10.02 3.92 |
| 50. | teams 6 0.04 3 0.00 + 15.04 3.92 |
| 51. | spina 4 0.03 2 0.00 + 10.02 3.92 |
| 52. | reassured 4 0.03 2 0.00 + 10.02 3.92 |
| 53. | private 6 0.04 3 0.00 + 15.04 3.92 |
| 54. | notes 4 0.03 2 0.00 + 10.02 3.92 |
| 55. | bifida 4 0.03 2 0.00 + 10.02 3.92 |
| 56. | appreciate 4 0.03 2 0.00 + 10.02 3.92 |
| 57. | you 38 0.26 21 0.02 + 91.47 3.77 |
| 58. | palliative 7 0.05 4 0.00 + 16.61 3.72 |
| 59. | memory 5 0.03 3 0.00 + 11.62 3.65 |
| 60. | often 16 0.11 10 0.01 + 36.50 3.59 |
| 61. | nor 8 0.06 5 0.00 + 18.25 3.59 |
| 62. | my 11 0.08 7 0.01 + 24.89 3.57 |
| 63. | certification 11 0.08 7 0.01 + 24.89 3.57 |
| 64. | helpful 20 0.14 13 0.01 + 44.81 3.54 |
| 65. | ventriculomegaly3 0.02 2 0.00 + 6.64 3.50 |
| 66. | unwanted 3 0.02 2 0.00 + 6.64 3.50 |
| 67. | steps 3 0.02 2 0.00 + 6.64 3.50 |
| 68. | specialised 3 0.02 2 0.00 + 6.64 3.50 |
| 69. | scheduled 3 0.02 2 0.00 + 6.64 3.50 |
| 70. | recovery 6 0.04 4 0.00 + 13.28 3.50 |
| 71. | mortem 3 0.02 2 0.00 + 6.64 3.50 |
| 72. | legally 3 0.02 2 0.00 + 6.64 3.50 |
| 73. | funeral 3 0.02 2 0.00 + 6.64 3.50 |
| 74. | ending 3 0.02 2 0.00 + 6.64 3.50 |
| 75. | confirmation 3 0.02 2 0.00 + 6.64 3.50 |
| 76. | communicating 3 0.02 2 0.00 + 6.64 3.50 |
| 77. | certificates 3 0.02 2 0.00 + 6.64 3.50 |
| 78. | anxious 3 0.02 2 0.00 + 6.64 3.50 |
| 79. | unexpected 10 0.07 7 0.01 + 21.62 3.43 |
| 80. | amniocentesis 10 0.07 7 0.01 + 21.62 3.43 |
| 81. | wording 4 0.03 3 0.00 + 8.35 3.33 |
| 82. | template 4 0.03 3 0.00 + 8.35 3.33 |
| 83. | survive 4 0.03 3 0.00 + 8.35 3.33 |
| 84. | prefer 4 0.03 3 0.00 + 8.35 3.33 |
| 85. | issue 4 0.03 3 0.00 + 8.35 3.33 |
| 86. | independent 8 0.06 6 0.01 + 16.70 3.33 |
| 87. | face 4 0.03 3 0.00 + 8.35 3.33 |
| 88. | afterwards 4 0.03 3 0.00 + 8.35 3.33 |
| 89. | team 21 0.15 16 0.01 + 43.48 3.31 |
| 90. | whatever 5 0.03 4 0.00 + 10.09 3.24 |
| 91. | our 10 0.07 8 0.01 + 20.17 3.24 |
| 92. | indicate 5 0.03 4 0.00 + 10.09 3.24 |
| 93. | consultation 5 0.03 4 0.00 + 10.09 3.24 |
| 94. | challenging 5 0.03 4 0.00 + 10.09 3.24 |
| 95. | anencephaly 5 0.03 4 0.00 + 10.09 3.24 |
| 96. | parents 198 1.38 162 0.15 + 394.52 3.21 |
| 97. | nuchal 6 0.04 5 0.00 + 11.83 3.18 |
| 98. | next 6 0.04 5 0.00 + 11.83 3.18 |
| 99. | burial 6 0.04 5 0.00 + 11.83 3.18 |
| 100. | certificate 15 0.10 13 0.01 + 28.93 3.12 |
| 101. | nhs 39 0.27 34 0.03 + 74.96 3.11 |
| 102. | end 8 0.06 7 0.01 + 15.34 3.11 |
| 103. | appointment 17 0.12 15 0.01 + 32.45 3.10 |
| 104. | expect 11 0.08 10 0.01 + 20.63 3.05 |
| 105. | finding 12 0.08 11 0.01 + 22.39 3.04 |
| 106. | will 152 1.06 143 0.13 + 279.17 3.00 |
| 107. | condition 35 0.24 34 0.03 + 63.02 2.96 |
| 108. | sands 5 0.03 5 0.00 + 8.84 2.92 |
| 109. | personal 4 0.03 4 0.00 + 7.07 2.92 |
| 110. | molar 4 0.03 4 0.00 + 7.07 2.92 |
| 111. | limitations 5 0.03 5 0.00 + 8.84 2.92 |
| 112. | leaflets 4 0.03 4 0.00 + 7.07 2.92 |
| 113. | isolated 5 0.03 5 0.00 + 8.84 2.92 |
| 114. | individualised 8 0.06 8 0.01 + 14.14 2.92 |
| 115. | extra 7 0.05 7 0.01 + 12.37 2.92 |
| 116. | depend 4 0.03 4 0.00 + 7.07 2.92 |
| 117. | cleft 5 0.03 5 0.00 + 8.84 2.92 |
| 118. | we 34 0.24 35 0.03 + 58.97 2.87 |
| 119. | through 14 0.10 15 0.01 + 23.64 2.82 |
| 120. | sensitively 7 0.05 8 0.01 + 11.30 2.72 |
| 121. | find 14 0.10 16 0.01 + 22.60 2.72 |
| 122. | distressing 7 0.05 8 0.01 + 11.30 2.72 |
| 123. | trust 6 0.04 7 0.01 + 9.54 2.69 |
| 124. | decide 12 0.08 14 0.01 + 19.09 2.69 |
| 125. | wait 5 0.03 6 0.01 + 7.79 2.65 |
| 126. | look 5 0.03 6 0.01 + 7.79 2.65 |
| 127. | explained 5 0.03 6 0.01 + 7.79 2.65 |
| 128. | documented 5 0.03 6 0.01 + 7.79 2.65 |
| 129. | discuss 21 0.15 26 0.02 + 31.96 2.61 |
| 130. | sensitive 12 0.08 15 0.01 + 18.13 2.59 |
| 131. | involve 8 0.06 10 0.01 + 12.09 2.59 |
| 132. | take 21 0.15 27 0.02 + 31.05 2.55 |
| 133. | medicine 35 0.24 45 0.04 + 51.74 2.55 |
| 134. | parent 6 0.04 8 0.01 + 8.62 2.50 |
| 135. | continuity 6 0.04 8 0.01 + 8.62 2.50 |
| 136. | key 25 0.17 34 0.03 + 35.33 2.47 |
| 137. | down 8 0.06 11 0.01 + 11.20 2.46 |
| 138. | testing 37 0.26 51 0.05 + 51.71 2.45 |
| 139. | renal 5 0.03 7 0.01 + 6.90 2.43 |
| 140. | raised 5 0.03 7 0.01 + 6.90 2.43 |
| 141. | opportunity 10 0.07 14 0.01 + 13.80 2.43 |
| 142. | patients 12 0.08 17 0.02 + 16.39 2.41 |
| 143. | choices 7 0.05 10 0.01 + 9.49 2.40 |
| 144. | difficult 18 0.13 26 0.02 + 24.18 2.39 |
| 145. | vary 6 0.04 9 0.01 + 7.80 2.33 |
| 146. | third 8 0.06 12 0.01 + 10.40 2.33 |
| 147. | open 6 0.04 9 0.01 + 7.80 2.33 |
| 148. | possibility 11 0.08 17 0.02 + 13.91 2.29 |
| 149. | hospital 16 0.11 25 0.02 + 20.04 2.27 |
| 150. | themselves 7 0.05 11 0.01 + 8.72 2.26 |

**Table 4: Negative keywords in Charity-HP sub-corpus (12,162 words) compared to Medical-HP sub-corpus (91,437 words)**

Excluded zeros in column 1, min. freq 1, LL greater than 6.63, sorted by effect size (LogRatio, cut off at -1), all results:

**Item	O1	%1	O2	%2	LL	LogRatio**

| 1. | recommendation 1 0.01 340 0.31 - 75.24 -5.49 |
| --- | --- |
| 2. | recurrent 1 0.01 214 0.20 - 44.81 -4.83 |
| 3. | late 1 0.01 199 0.18 - 41.22 -4.72 |
| 4. | studies 1 0.01 144 0.13 - 28.18 -4.25 |
| 5. | perinatal 1 0.01 107 0.10 - 19.56 -3.83 |
| 6. | recommendations 2 0.01 215 0.20 - 39.36 -3.83 |
| 7. | twin 1 0.01 107 0.10 - 19.56 -3.83 |
| 8. | shown 1 0.01 102 0.09 - 18.41 -3.76 |
| 9. | abnormalities 1 0.01 101 0.09 - 18.18 -3.74 |
| 10. | evidence 5 0.03 486 0.45 - 86.58 -3.69 |
| 11. | maternal 1 0.01 96 0.09 - 17.04 -3.67 |
| 12. | 2012 1 0.01 92 0.08 - 16.13 -3.61 |
| 13. | uterine 1 0.01 89 0.08 - 15.45 -3.56 |
| 14. | rationale 1 0.01 84 0.08 - 14.32 -3.48 |
| 15. | live 1 0.01 83 0.08 - 14.10 -3.46 |
| 16. | preterm 1 0.01 81 0.07 - 13.65 -3.42 |
| 17. | cervical 1 0.01 74 0.07 - 12.08 -3.29 |
| 18. | nice 1 0.01 74 0.07 - 12.08 -3.29 |
| 19. | history 1 0.01 71 0.07 - 11.42 -3.23 |
| 20. | &lsqb; 5 0.03 330 0.30 - 51.60 -3.13 |
| 21. | examination 1 0.01 66 0.06 - 10.32 -3.13 |
| 22. | &rsqb; 5 0.03 329 0.30 - 51.39 -3.12 |
| 23. | ? 6 0.04 392 0.36 - 61.05 -3.11 |
| 24. | although 1 0.01 64 0.06 - 9.88 -3.08 |
| 25. | assessment 2 0.01 125 0.12 - 19.12 -3.05 |
| 26. | 4 2 0.01 124 0.11 - 18.90 -3.04 |
| 27. | gestational 2 0.01 123 0.11 - 18.68 -3.03 |
| 28. | associated 2 0.01 119 0.11 - 17.82 -2.98 |
| 29. | 2023 1 0.01 59 0.05 - 8.80 -2.97 |
| 30. | future 1 0.01 59 0.05 - 8.80 -2.97 |
| 31. | treatment 4 0.03 217 0.20 - 31.14 -2.85 |
| 32. | she 1 0.01 51 0.05 - 7.10 -2.76 |
| 33. | common 1 0.01 49 0.05 - 6.68 -2.70 |
| 34. | level 5 0.03 246 0.23 - 33.61 -2.70 |
| 35. | 3 3 0.02 143 0.13 - 19.21 -2.66 |
| 36. | been 5 0.03 223 0.21 - 28.86 -2.56 |
| 37. | women 21 0.15 910 0.84 - 115.79 -2.52 |
| 38. | stillbirth 3 0.02 124 0.11 - 15.33 -2.45 |
| 39. | less 2 0.01 82 0.08 - 10.09 -2.44 |
| 40. | rate 2 0.01 82 0.08 - 10.09 -2.44 |
| 41. | death 3 0.02 122 0.11 - 14.93 -2.43 |
| 42. | number 2 0.01 81 0.07 - 9.88 -2.42 |
| 43. | practice 4 0.03 161 0.15 - 19.57 -2.41 |
| 44. | risk 8 0.06 313 0.29 - 37.35 -2.37 |
| 45. | 2 2 0.01 77 0.07 - 9.09 -2.35 |
| 46. | complications 2 0.01 76 0.07 - 8.89 -2.33 |
| 47. | woman 5 0.03 188 0.17 - 21.84 -2.32 |
| 48. | recommended 4 0.03 141 0.13 - 15.64 -2.22 |
| 49. | pregnancies 6 0.04 203 0.19 - 21.83 -2.16 |
| 50. | levels 2 0.01 67 0.06 - 7.15 -2.15 |
| 51. | ectopic 9 0.06 300 0.28 - 31.89 -2.14 |
| 52. | was 7 0.05 233 0.21 - 24.74 -2.14 |
| 53. | c 2 0.01 65 0.06 - 6.77 -2.11 |
| 54. | misoprostol 2 0.01 65 0.06 - 6.77 -2.11 |
| 55. | research 2 0.01 65 0.06 - 6.77 -2.11 |
| 56. | d 5 0.03 161 0.15 - 16.65 -2.09 |
| 57. | caesarean 3 0.02 92 0.08 - 9.13 -2.02 |
| 58. | second 3 0.02 92 0.08 - 9.13 -2.02 |
| 59. | age 3 0.02 86 0.08 - 8.04 -1.93 |
| 60. | used 4 0.03 113 0.10 - 10.42 -1.90 |
| 61. | review 4 0.03 109 0.10 - 9.70 -1.85 |
| 62. | ultrasound 10 0.07 257 0.24 - 21.55 -1.77 |
| 63. | ; 7 0.05 171 0.16 - 13.57 -1.69 |
| 64. | management 14 0.10 308 0.28 - 21.55 -1.54 |
| 65. | early 9 0.06 173 0.16 - 10.00 -1.35 |
| 66. | than 11 0.08 199 0.18 - 10.43 -1.26 |
| 67. | quality 8 0.06 144 0.13 - 7.48 -1.25 |
| 68. | were 10 0.07 172 0.16 - 8.25 -1.19 |
| 69. | miscarriage 30 0.21 477 0.44 - 19.61 -1.07 |

**Table 5: Positive keywords in Medical-P sub-corpus (28,931 words) compared to Charity-P sub-corpus (212,158 words)**

Excluded zeros in column 1, min. freq 1, LL greater than 6.63, sorted by effect size (LogRatio), Sample of top 150 results:

**Item	O1	%1	O2	%2	LL LogRatio**

| 1. | gtd 16 0.05 1 0.00 + 60.46 6.87 |
| --- | --- |
| 2. | raw 7 0.02 1 0.00 + 23.90 5.68 |
| 3. | obesity 6 0.02 1 0.00 + 19.94 5.46 |
| 4. | obese 5 0.01 1 0.00 + 16.04 5.19 |
| 5. | 'women' 5 0.01 1 0.00 + 16.04 5.19 |
| 6. | 'woman' 5 0.01 1 0.00 + 16.04 5.19 |
| 7. | reporting 4 0.01 1 0.00 + 12.21 4.87 |
| 8. | answering 4 0.01 1 0.00 + 12.21 4.87 |
| 9. | poultry 3 0.01 1 0.00 + 8.47 4.46 |
| 10. | jab 3 0.01 1 0.00 + 8.47 4.46 |
| 11. | foetal 3 0.01 1 0.00 + 8.47 4.46 |
| 12. | cooked 3 0.01 1 0.00 + 8.47 4.46 |
| 13. | childhood 3 0.01 1 0.00 + 8.47 4.46 |
| 14. | 250 3 0.01 1 0.00 + 8.47 4.46 |
| 15. | inclusive 5 0.01 2 0.00 + 13.33 4.19 |
| 16. | gender 5 0.01 2 0.00 + 13.33 4.19 |
| 17. | fruit 5 0.01 2 0.00 + 13.33 4.19 |
| 18. | bacterial 5 0.01 2 0.00 + 13.33 4.19 |
| 19. | weakened 6 0.02 3 0.00 + 14.74 3.87 |
| 20. | vegetables 5 0.01 3 0.00 + 11.37 3.61 |
| 21. | non-tubal 5 0.01 3 0.00 + 11.37 3.61 |
| 22. | meat 5 0.01 3 0.00 + 11.37 3.61 |
| 23. | flu 5 0.01 3 0.00 + 11.37 3.61 |
| 24. | rcog 21 0.06 13 0.01 + 47.10 3.56 |
| 25. | overweight 8 0.02 5 0.00 + 17.86 3.55 |
| 26. | pcos 6 0.02 4 0.00 + 12.99 3.46 |
| 27. | gtn 16 0.05 11 0.00 + 34.13 3.41 |
| 28. | foetus 10 0.03 7 0.00 + 21.14 3.39 |
| 29. | abortion 15 0.04 11 0.00 + 30.96 3.32 |
| 30. | vision 4 0.01 3 0.00 + 8.16 3.29 |
| 31. | trisomy 4 0.01 3 0.00 + 8.16 3.29 |
| 32. | illnesses 4 0.01 3 0.00 + 8.16 3.29 |
| 33. | &e); 4 0.01 3 0.00 + 8.16 3.29 |
| 34. | points 5 0.01 4 0.00 + 9.85 3.19 |
| 35. | identity 5 0.01 4 0.00 + 9.85 3.19 |
| 36. | confirmed 16 0.05 13 0.01 + 31.25 3.17 |
| 37. | suspected 11 0.03 9 0.00 + 21.40 3.16 |
| 38. | obstetric 7 0.02 6 0.00 + 13.26 3.10 |
| 39. | itching 7 0.02 6 0.00 + 13.26 3.10 |
| 40. | foods 16 0.05 15 0.01 + 28.71 2.97 |
| 41. | thrombophilia 4 0.01 4 0.00 + 6.89 2.87 |
| 42. | spread 5 0.01 5 0.00 + 8.61 2.87 |
| 43. | inserting 5 0.01 5 0.00 + 8.61 2.87 |
| 44. | headache 4 0.01 4 0.00 + 6.89 2.87 |
| 45. | examinations 4 0.01 4 0.00 + 6.89 2.87 |
| 46. | advisable 4 0.01 4 0.00 + 6.89 2.87 |
| 47. | abnormal 15 0.04 17 0.01 + 23.69 2.69 |
| 48. | removing 7 0.02 8 0.00 + 10.99 2.68 |
| 49. | one-off 7 0.02 8 0.00 + 10.99 2.68 |
| 50. | hands 6 0.02 7 0.00 + 9.28 2.65 |
| 51. | heparin 5 0.01 6 0.00 + 7.57 2.61 |
| 52. | delay 5 0.01 6 0.00 + 7.57 2.61 |
| 53. | eat 11 0.03 14 0.01 + 15.91 2.52 |
| 54. | shape 7 0.02 9 0.00 + 10.04 2.51 |
| 55. | relative 7 0.02 9 0.00 + 10.04 2.51 |
| 56. | clotting 7 0.02 9 0.00 + 10.04 2.51 |
| 57. | 'll 38 0.11 50 0.02 + 53.50 2.48 |
| 58. | recovering 9 0.03 12 0.00 + 12.53 2.46 |
| 59. | followed 6 0.02 8 0.00 + 8.36 2.46 |
| 60. | cholestasis 9 0.03 12 0.00 + 12.53 2.46 |
| 61. | operation 56 0.16 77 0.03 + 76.01 2.41 |
| 62. | inherited 8 0.02 11 0.00 + 10.86 2.41 |
| 63. | tampons 5 0.01 7 0.00 + 6.68 2.39 |
| 64. | drinking 10 0.03 14 0.01 + 13.37 2.39 |
| 65. | 35 5 0.01 7 0.00 + 6.68 2.39 |
| 66. | smoking 22 0.06 31 0.01 + 29.24 2.38 |
| 67. | wishes 7 0.02 10 0.00 + 9.19 2.36 |
| 68. | wish 24 0.07 35 0.01 + 30.95 2.33 |
| 69. | drugs 13 0.04 19 0.01 + 16.73 2.33 |
| 70. | carried 22 0.06 32 0.01 + 28.44 2.33 |
| 71. | uncommon 6 0.02 9 0.00 + 7.54 2.29 |
| 72. | smoke 6 0.02 9 0.00 + 7.54 2.29 |
| 73. | combination 6 0.02 9 0.00 + 7.54 2.29 |
| 74. | causing 8 0.02 12 0.00 + 10.06 2.29 |
| 75. | becoming 6 0.02 9 0.00 + 7.54 2.29 |
| 76. | terms 22 0.06 34 0.01 + 26.90 2.24 |
| 77. | reduced 7 0.02 11 0.00 + 8.43 2.22 |
| 78. | tummy 20 0.06 32 0.01 + 23.66 2.19 |
| 79. | weight 16 0.05 26 0.01 + 18.65 2.17 |
| 80. | preventing 6 0.02 10 0.00 + 6.82 2.14 |
| 81. | alcohol 15 0.04 25 0.01 + 17.05 2.14 |
| 82. | involves 16 0.05 27 0.01 + 17.96 2.12 |
| 83. | probe 7 0.02 12 0.00 + 7.73 2.10 |
| 84. | discuss 36 0.10 62 0.02 + 39.57 2.09 |
| 85. | diet 9 0.03 16 0.01 + 9.57 2.04 |
| 86. | medicine 30 0.08 54 0.02 + 31.48 2.02 |
| 87. | food 11 0.03 20 0.01 + 11.42 2.01 |
| 88. | tubes 12 0.03 22 0.01 + 12.34 2.00 |
| 89. | laparoscopy 12 0.03 22 0.01 + 12.34 2.00 |
| 90. | asleep 7 0.02 13 0.01 + 7.10 1.98 |
| 91. | movements 15 0.04 28 0.01 + 15.12 1.97 |
| 92. | drink 9 0.03 17 0.01 + 8.95 1.96 |
| 93. | affects 9 0.03 17 0.01 + 8.95 1.96 |
| 94. | confirm 11 0.03 21 0.01 + 10.81 1.94 |
| 95. | naturally 14 0.04 27 0.01 + 13.60 1.93 |
| 96. | vaginal 43 0.12 83 0.03 + 41.74 1.92 |
| 97. | serious 19 0.05 37 0.01 + 18.26 1.91 |
| 98. | pre-eclampsia 13 0.04 26 0.01 + 12.10 1.87 |
| 99. | harm 8 0.02 16 0.01 + 7.45 1.87 |
| 100. | further 43 0.12 86 0.03 + 40.04 1.87 |
| 101. | choices 16 0.05 32 0.01 + 14.90 1.87 |
| 102. | 48 10 0.03 20 0.01 + 9.31 1.87 |
| 103. | advised 20 0.06 41 0.02 + 18.08 1.84 |
| 104. | infections 12 0.03 25 0.01 + 10.63 1.81 |
| 105. | eating 10 0.03 21 0.01 + 8.77 1.80 |
| 106. | 100 20 0.06 42 0.02 + 17.55 1.80 |
| 107. | urine 14 0.04 30 0.01 + 11.97 1.77 |
| 108. | contraception 14 0.04 30 0.01 + 11.97 1.77 |
| 109. | remove 22 0.06 48 0.02 + 18.38 1.75 |
| 110. | end 20 0.06 44 0.02 + 16.53 1.74 |
| 111. | transvaginal 9 0.03 20 0.01 + 7.34 1.72 |
| 112. | discharge 13 0.04 29 0.01 + 10.55 1.72 |
| 113. | association 13 0.04 29 0.01 + 10.55 1.72 |
| 114. | follow-up 22 0.06 50 0.02 + 17.41 1.69 |
| 115. | chemotherapy 11 0.03 25 0.01 + 8.70 1.69 |
| 116. | healthcare 57 0.16 131 0.05 + 44.42 1.67 |
| 117. | chances 14 0.04 33 0.01 + 10.53 1.64 |
| 118. | sperm 10 0.03 24 0.01 + 7.33 1.61 |
| 119. | appointments 16 0.05 39 0.02 + 11.47 1.59 |
| 120. | options 50 0.14 123 0.05 + 35.36 1.57 |
| 121. | thyroid 10 0.03 25 0.01 + 6.90 1.55 |
| 122. | recommended 14 0.04 35 0.01 + 9.66 1.55 |
| 123. | assessment 12 0.03 30 0.01 + 8.28 1.55 |
| 124. | molar 80 0.23 201 0.08 + 54.81 1.54 |
| 125. | antenatal 19 0.05 48 0.02 + 12.91 1.54 |
| 126. | depend 13 0.04 33 0.01 + 8.77 1.53 |
| 127. | sign 16 0.05 41 0.02 + 10.64 1.52 |
| 128. | become 20 0.06 53 0.02 + 12.61 1.47 |
| 129. | 111 11 0.03 29 0.01 + 6.99 1.47 |
| 130. | mother 21 0.06 56 0.02 + 13.10 1.46 |
| 131. | tissue 35 0.10 94 0.04 + 21.58 1.45 |
| 132. | hormone 24 0.07 65 0.03 + 14.60 1.44 |
| 133. | develop 27 0.08 73 0.03 + 16.47 1.44 |
| 134. | infection 33 0.09 90 0.03 + 19.84 1.43 |
| 135. | 3 34 0.10 93 0.04 + 20.34 1.42 |
| 136. | caused 20 0.06 56 0.02 + 11.50 1.39 |
| 137. | situation 19 0.05 54 0.02 + 10.64 1.37 |
| 138. | severe 19 0.05 54 0.02 + 10.64 1.37 |
| 139. | fallopian 35 0.10 99 0.04 + 19.77 1.37 |
| 140. | known 29 0.08 83 0.03 + 16.04 1.36 |
| 141. | complete 14 0.04 40 0.02 + 7.77 1.36 |
| 142. | immediately 17 0.05 49 0.02 + 9.29 1.35 |
| 143. | : 155 0.44 446 0.17 + 84.93 1.35 |
| 144. | increase 26 0.07 76 0.03 + 13.85 1.33 |
| 145. | advice 36 0.10 105 0.04 + 19.25 1.33 |
| 146. | uterus 44 0.12 129 0.05 + 23.31 1.32 |
| 147. | usually 82 0.23 242 0.09 + 42.91 1.31 |
| 148. | avoid 23 0.06 69 0.03 + 11.67 1.29 |
| 149. | reduce 22 0.06 67 0.03 + 10.85 1.27 |
| 150. | offered 62 0.18 188 0.07 + 30.83 1.27 |

**Table 6: Negative keywords in Medical-P sub-corpus (28,931 words) compared to Charity-P sub-corpus (212,158 words)**

Excluded zeros in column 1, min. freq 1, LL greater than 6.63, sorted by effect size (LogRatio, cut off at -1), all results:

**Item	O1	%1	O2	%2	LL LogRatio**

| 1. | our 1 0.00 467 0.18 - 109.48 -5.99 |
| --- | --- |
| 2. | us 1 0.00 206 0.08 - 44.31 -4.81 |
| 3. | say 1 0.00 138 0.05 - 27.70 -4.24 |
| 4. | &quot; 4 0.01 501 0.19 - 98.52 -4.10 |
| 5. | partners 1 0.00 85 0.03 - 15.10 -3.54 |
| 6. | perhaps 1 0.00 82 0.03 - 14.40 -3.48 |
| 7. | everyone 1 0.00 76 0.03 - 13.02 -3.38 |
| 8. | was 7 0.02 476 0.18 - 78.34 -3.21 |
| 9. | fetal 1 0.00 65 0.03 - 10.51 -3.15 |
| 10. | helped 1 0.00 62 0.02 - 9.84 -3.08 |
| 11. | doctors 2 0.01 122 0.05 - 19.23 -3.06 |
| 12. | mental 2 0.01 119 0.05 - 18.56 -3.02 |
| 13. | book 1 0.00 59 0.02 - 9.17 -3.01 |
| 14. | did 2 0.01 117 0.05 - 18.12 -3.00 |
| 15. | upsetting 1 0.00 57 0.02 - 8.73 -2.96 |
| 16. | understanding 1 0.00 54 0.02 - 8.06 -2.88 |
| 17. | leave 2 0.01 107 0.04 - 15.91 -2.87 |
| 18. | me 3 0.01 157 0.06 - 23.10 -2.84 |
| 19. | describe 1 0.00 52 0.02 - 7.63 -2.83 |
| 20. | emotions 2 0.01 104 0.04 - 15.25 -2.83 |
| 21. | here 5 0.01 256 0.10 - 37.27 -2.81 |
| 22. | carry 1 0.00 51 0.02 - 7.41 -2.80 |
| 23. | nipt 1 0.00 51 0.02 - 7.41 -2.80 |
| 24. | space 1 0.00 51 0.02 - 7.41 -2.80 |
| 25. | report 1 0.00 50 0.02 - 7.19 -2.77 |
| 26. | thing 1 0.00 50 0.02 - 7.19 -2.77 |
| 27. | caring 1 0.00 49 0.02 - 6.98 -2.74 |
| 28. | hear 1 0.00 49 0.02 - 6.98 -2.74 |
| 29. | upset 1 0.00 49 0.02 - 6.98 -2.74 |
| 30. | process 3 0.01 142 0.05 - 19.86 -2.69 |
| 31. | hard 3 0.01 140 0.05 - 19.43 -2.67 |
| 32. | children 3 0.01 139 0.05 - 19.22 -2.66 |
| 33. | cope 2 0.01 91 0.04 - 12.46 -2.64 |
| 34. | loss 19 0.05 828 0.32 - 110.61 -2.57 |
| 35. | new 2 0.01 87 0.03 - 11.61 -2.57 |
| 36. | progesterone 2 0.01 80 0.03 - 10.15 -2.45 |
| 37. | experiences 2 0.01 79 0.03 - 9.94 -2.43 |
| 38. | place 3 0.01 117 0.05 - 14.61 -2.41 |
| 39. | black 2 0.01 73 0.03 - 8.72 -2.32 |
| 40. | memory 2 0.01 73 0.03 - 8.72 -2.32 |
| 41. | result 3 0.01 106 0.04 - 12.38 -2.27 |
| 42. | share 2 0.01 69 0.03 - 7.92 -2.24 |
| 43. | staff 3 0.01 98 0.04 - 10.79 -2.16 |
| 44. | them 17 0.05 545 0.21 - 59.13 -2.13 |
| 45. | her 6 0.02 189 0.07 - 20.22 -2.10 |
| 46. | i 27 0.08 791 0.31 - 79.62 -2.00 |
| 47. | late 3 0.01 88 0.03 - 8.87 -2.00 |
| 48. | part 4 0.01 114 0.04 - 11.20 -1.96 |
| 49. | might 24 0.07 673 0.26 - 65.13 -1.94 |
| 50. | those 3 0.01 82 0.03 - 7.75 -1.90 |
| 51. | ' 5 0.01 133 0.05 - 12.24 -1.86 |
| 52. | remember 5 0.01 130 0.05 - 11.70 -1.83 |
| 53. | please 5 0.01 128 0.05 - 11.34 -1.81 |
| 54. | tell 6 0.02 154 0.06 - 13.68 -1.81 |
| 55. | happened 6 0.02 151 0.06 - 13.14 -1.78 |
| 56. | just 7 0.02 176 0.07 - 15.30 -1.78 |
| 57. | feelings 10 0.03 249 0.10 - 21.43 -1.77 |
| 58. | were 7 0.02 165 0.06 - 13.37 -1.69 |
| 59. | yourself 7 0.02 161 0.06 - 12.68 -1.65 |
| 60. | we 33 0.09 752 0.29 - 58.60 -1.64 |
| 61. | life 5 0.01 112 0.04 - 8.55 -1.61 |
| 62. | parents 15 0.04 333 0.13 - 25.15 -1.60 |
| 63. | research 6 0.02 133 0.05 - 10.02 -1.60 |
| 64. | grief 10 0.03 221 0.09 - 16.60 -1.59 |
| 65. | physical 6 0.02 132 0.05 - 9.86 -1.59 |
| 66. | through 20 0.06 430 0.17 - 31.20 -1.55 |
| 67. | funeral 8 0.02 170 0.07 - 12.15 -1.54 |
| 68. | know 17 0.05 356 0.14 - 24.96 -1.52 |
| 69. | person 5 0.01 101 0.04 - 6.74 -1.46 |
| 70. | would 12 0.03 238 0.09 - 15.49 -1.44 |
| 71. | look 9 0.03 176 0.07 - 11.22 -1.42 |
| 72. | things 14 0.04 272 0.10 - 17.18 -1.41 |
| 73. | people 33 0.09 632 0.24 - 39.09 -1.39 |
| 74. | babies 8 0.02 152 0.06 - 9.29 -1.38 |
| 75. | their 34 0.10 638 0.25 - 38.26 -1.36 |
| 76. | again 18 0.05 313 0.12 - 16.58 -1.25 |
| 77. | my 17 0.05 285 0.11 - 14.15 -1.19 |
| 78. | even 15 0.04 247 0.10 - 11.87 -1.17 |
| 79. | second 10 0.03 164 0.06 - 7.82 -1.16 |
| 80. | like 19 0.05 309 0.12 - 14.50 -1.15 |
| 81. | offer 10 0.03 163 0.06 - 7.68 -1.15 |
| 82. | helpful 16 0.05 259 0.10 - 12.05 -1.14 |
| 83. | post-mortem 11 0.03 178 0.07 - 8.27 -1.14 |
| 84. | own 10 0.03 160 0.06 - 7.27 -1.13 |
| 85. | feel 48 0.14 765 0.30 - 34.51 -1.12 |
| 86. | others 11 0.03 170 0.07 - 7.21 -1.08 |
| 87. | difficult 18 0.05 276 0.11 - 11.51 -1.07 |
| 88. | right 13 0.04 197 0.08 - 8.01 -1.05 |
| 89. | they 71 0.20 1060 0.41 - 41.72 -1.03 |
| 90. | n't 42 0.12 622 0.24 - 24.05 -1.02 |
| 91. | ask 17 0.05 250 0.10 - 9.51 -1.01 |
